# The phenotypic and genetic relationship between adolescent mental health and time spent on social media, gaming, and TV

**DOI:** 10.1101/2023.09.14.23295537

**Authors:** Evgeniia Frei, Oleksandr Frei, Piotr Jaholkowski, Nadine Parker, Pravesh Parekh, Alexey A. Shadrin, Espen Hagen, Nora R. Bakken, Viktoria Birkenæs, Helga Ask, Ole A. Andreassen, Olav B. Smeland

## Abstract

**Background:** Screen use is linked to adverse mental health outcomes in adolescents, but differences across psychiatric diagnoses, mental symptoms, and types of screen-based activities remain unclear. Moreover, the extent to which genetic factors contribute to these associations is largely unknown.

**Methods:** We analysed longitudinal data from 23,790 adolescents (14-16 years of age) in the Norwegian Mother, Father, and Child Cohort Study (MoBa), linking questionnaire responses on screen use to registry-based psychiatric diagnoses and self-reported mental symptoms. Using regression models, we assessed associations between mental health outcomes and gaming, social media use, and TV watching. We also evaluated whether genetic liability to major psychiatric disorders, as indexed by polygenic risk scores (PRSs), was associated with screen use, and estimated the degree of genetic confounding using genetic sensitivity analyses.

**Findings:** Spending 3–4 hours/day or more on any screen-based activity was associated with increased odds of a lifetime diagnosis of depressive, anxiety/stress-related, and hyperkinetic disorders. Similarly, minimal social media engagement was associated with increased odds of any psychiatric diagnosis, most strongly with pervasive developmental disorders. Associations with mental symptoms closely followed those observed for clinical diagnoses. The PRSs of major depression, anorexia nervosa, attention-deficit hyperactivity disorder, and autism spectrum disorder were significantly associated with screen use, suggesting shared genetic liability. Genetic sensitivity analyses indicated that a substantial proportion of the observed phenotypic associations may be attributable to genetic factors.

**Interpretation:** Our findings suggest a complex, bidirectional relationship between screen use and adolescent mental health, across both mental disorders and traits. Both high and low screen use may reflect underlying mental health vulnerabilities. The associations appear partly driven by shared genetic susceptibility, underscoring the importance of accounting for individual risk. These results support the need for individualized, context-sensitive digital media use guidelines.

**Funding:** Research Council of Norway (Grant N°324499).

## Research in context

### Evidence before this study

A growing body of research has indicated an association between time spent using screen devices and adverse mental health outcomes among adolescents. Yet, the extent to which these associations differ across mental disorders and screen-based activities remain unclear. Although the evidence is sparse, some countries are implementing or considering strict screen device restrictions for adolescents. In February 2023, we searched PubMed for studies published in English, using a combination of the terms “screen time”, “screen”, “digital media”, “smartphone”, “social media”, “internet”, “television”, “video”, “gaming”, “mental health”, “mental disorders”, “psychiatric disorders”, “anxiety”, “psychological wellbeing”, “internalizing symptoms”, “behavioral problems”; the search was last updated in March 2025. Most studies report significant associations between more screen time and various aspects of adolescent mental health, while a few find no adverse links. Furthermore, few studies were linked to clinical diagnoses from health registries, while the range of mental health outcomes was often narrow. Additionally, most studies focused on either overall screen time or a single type of screen-based activity, limiting comparability across screen domains. Genetic studies have demonstrated that screen-based behaviours are heritable traits. At least two studies have shown that genetic confounding may partly explain associations between screen time and mental health problems; however, these studies examined a limited scope of mental health outcomes and lacked linkage to clinical data.

### Added value of this study

Here we provide a comprehensive investigation of the associations between adolescent mental health and screen-based behaviours (gaming, social media use, and TV watching) across diagnosis and mental traits. We leveraged a large, genotyped population-based cohort, the Norwegian Mother, Father, and Child Cohort Study (MoBa), with self-report questionnaires on screen use and mental health symptoms as well as linkage to clinically diagnosed mental disorders. We examined these associations at both the phenotypic and genetic level, incorporating polygenic risk scores and genetic sensitivity analyses to evaluate the role of shared genetic liability and potential genetic confounding.

High levels of gaming, social media use, and TV watching were associated with increased odds of a diagnosis of depressive, anxiety/stress-related, and hyperkinetic disorders, which closely mirrored patterns observed for mental symptoms in healthy adolescents. A U-shaped pattern emerged with minimal social media engagement was also linked to higher odds of any psychiatric diagnosis, potentially reflecting impaired social functioning seen across mental disorders, and particularly neurodevelopmental disorders, which showed the strongest association with the lowest social media use.

The phenotypic and genetic findings were generally aligned. Polygenic risk scores for major depression, anorexia nervosa, attention-deficit hyperactivity disorder, and autism spectrum disorder were significantly associated with screen use, suggesting shared genetic liability. Moreover, associations between moderate-to-high screen use and most diagnoses were substantially confounded by genetic factors, underscoring the importance of integrating genetic perspective in the study of complex behavioural phenotypes in the context of psychiatry.

### Implications of all the available evidence

Our findings suggest a bidirectional relationship between screen time and adolescent mental health, with implications for policymakers. While the patterns of association differ across disorders and screen activities, excessive screen time was consistently associated with adverse mental health outcomes across multiple domains. Our genetic findings indicate that shared genetic factors may contribute to the co-occurrence of excessive screen usage and mental illness, highlighting that a subgroup may be at elevated risk for both. The U-shaped pattern with strong associations between minimal social media use and mental illness may reflect negative peer dynamics, pre-existing/pre-morbid mental health conditions or impaired social functioning, indicating that very low social media use may serve as a marker of vulnerability. Overall, our findings underscore the importance of considering individual vulnerability when assessing the mental health impact of screen use among adolescents. Genetically informed designs and longitudinal datasets will be critical for disentangling causal relationships.

## Background

The availability of digital devices continues to increase, and adolescents spend more time on screen-based activities than ever before. Across European countries, most children aged 9-16 use their smartphones several times each day and spend up to 4-5 hours/day online.^1^ As the time devoted to digital devices has risen, so too have the concerns about the relationship of screen behaviors with youth’s mental wellbeing.

Sound mental health is of particular importance during adolescence, as this is a critical and formative period when individuals begin their transition to adulthood. Adolescents with mental health problems have lower quality of life, and are vulnerable to social exclusion, educational difficulties, and stigma.^2^ In recent decades, the prevalence of adolescent mental health problems has risen markedly,^3^ and mental disorders are becoming major contributors to health-related disability in children and youth.^4^

A growing body of research has suggested the potential benefits and harms of screen use for adolescent mental health. According to the recent reviews, most studies report significant associations between screen time and various problems of adolescent mental health, while only a few find no adverse links.^5–7^ Importantly, associations between screen use and mental health seems to vary by the specific type of mental health problem, with most significant links to internalizing symptoms like depression and anxiety.^8^ Associations with mental challenges seems to differ across types of screen activities, with social media showing more consistent negative effects.^6^ Moreover, the link between screen use and adolescent mental health can vary depending on the amount of time spent on screen use, with little effects associated with moderate use, while excessive use is more strongly associated with mental health problems.^9^ Finally, most studies rely on self-reported mental health measures and lack clinical information and data on psychiatric diagnoses. Thus, it has been difficult to translate the findings to clinical settings.

The fact that mental disorders and screen behaviours are heritable traits, highlights the importance of genetically informed designs in studying their association. While some studies have attempted to investigate the role of shared genetic factors linking screen time and mental health problems in young populations, the findings are unclear. Associations have been observed between polygenic risk scores (PRSs) of ADHD and depression and screen time in children aged 9-11 years.^10,11^ Moreover, the association between screen time and attention problems, as well as internalizing problems, was confounded by genetic factors^10^ Another study also found genetic influences on media use, although it focused on young adults rather than adolescents, and assessed general media use rather than specific screen activities.^12^ Overall, the potential of shared genetic factors between screen time and mental health problems in adolescents remains unknown and would be relevant to inform prevention measures.

Based on existing evidence linking screen use to adolescent mental health, we investigated whether these associations vary across different types of screen activities, are specific to certain mental disorders or mental traits, while incorporating both self-reported mental symptoms and psychiatric diagnoses. We used data from the Norwegian Mother, Father, and Child Cohort Study (MoBa).^13^ Additionally, we investigated the relationship between screen time and genetic liability to major psychiatric disorders, as indicated by PRSs derived from recent genome-wide association studies on schizophrenia (SCZ), bipolar disorder (BD), major depression (MD), autism spectrum disorder (ASD), attention-deficit hyperactivity disorder (ADHD), anxiety (ANX), anorexia nervosa (AN), and alcohol use disorder (AUD). Finally, we aimed to assess the role of genetic factors in the phenotypic associations.

## Methods

### Study sample

MoBa is a population-based pregnancy cohort study conducted by the Norwegian Institute of Public Health.^13^ Participants were recruited from all over Norway from 1999-2008. The women consented to participation in 41% of the pregnancies. The cohort includes approximately 114,500 children, 95,200 mothers, and 75,200 fathers. Blood samples obtained from the children’s umbilical cord at birth were used for genotyping.^14^ The study sample was restricted to adolescents (14-16 years of age) who answered the MoBa questionnaire Q-14year, with data linked to the Medical Birth Registry of Norway (MBRN) (*n* = 25,079). The MBRN is a national birth registry containing information about all births in Norway.

The establishment of MoBa and initial data collection was based on a license from the Norwegian Data Protection Agency and approval from The Regional Committees for Medical and Health Research Ethics. The MoBa cohort is currently regulated by the Norwegian Health Registry Act. The current study was approved by The Regional Committees for Medical and Health Research Ethics (2016/1226/REK Sør-Øst C) in Norway.

### Study variables

Each screen-based behaviour – television viewing, gaming, and social media use – was measured with one separate item (Table 1). For each item, participants rated the daily time spent on the activity from 1 to 6 (least to more time). Parental highest level of education was used as a proxy variable characterizing socioeconomic status (SES) (see Supplementary Text 1, Table S1 for details).

**Table 1.** Basic sociodemographic characteristics of the study sample (*n* = 23,790), and descriptive information on key study variables.

| **Basic demographic characteristics of the primary study sample** |  |  |
| --- | --- | --- |
| Age when questionnaire was answered, years, mean (SD) |  | 14.41 (0.51) |
| Sex assigned at birth, $n$ (%) | | |
| Male |  | 10,917 (45.89%) |
| Female |  | 12,873 (54.11%) |
| Gender identity, $n$ (%) | | |
| Boy |  | 10,591 (44.52%) |
| Girl |  | 12,412 (52.17%) |
| Trans person |  | 59 (0.25%) |
| Do not know / Do not wish to answer |  | 337 (1.42%) |
| NA |  | 391 (1.64%) |
| Socioeconomic status<sup>1</sup>, $n$ (%) | | |
| Low |  | 18,456 (77.58%) |
| High |  | 5009 (21.06%) |
| NA |  | 325 (1.37%) |
| **Descriptive information on key study variables** |  |  |
| Questions | Response options | $n$ (%) |
| How much time do you usually spend during one weekday on the following activities? |  |  |
| Watch movies/series/TV | 1 - Never/ rarely | 2189 (9.20%) |
|  | 2 - Less than 1 hour | 6227 (26.17%) |
|  | 3 - 1-2 hours | 10,374 (43.61%) |
|  | 4 - 3-4 hours | 3772 (15.86%) |
|  | 5 - 5-6 hours | 752 (3.16%) |
|  | 6 - 7 hours or more | 278 (1.17%) |
|  | NA | 198 (0.83%) |
| Playing games (on PC, TV, tablet, mobile etc.) | 1 - Never/ rarely | 5874 (24.69%) |
|  | 2 - Less than 1 hour | 6048 (25.42%) |
|  | 3 - 1-2 hours | 5920 (24.88%) |
|  | 4 - 3-4 hours | 3949 (16.60%) |
|  | 5 - 5-6 hours | 1253 (5.27%) |
|  | 6 - 7 hours or more | 507 (2.13%) |
|  | NA | 239 (1.00%) |
| Communicating with friends on social media | 1 - Never/ rarely | 756 (3.18%) |
|  | 2 - Less than 1 hour | 5134 (21.58%) |
|  | 3 - 1-2 hours | 8080 (33.96%) |
|  | 4 - 3-4 hours | 5950 (25.01%) |
|  | 5 - 5-6 hours | 2395 (10.07%) |
|  | 6 - 7 hours or more | 1301 (5.47%) |
|  | NA | 174 (0.73%) |

NA indicates missing values.
<sup>1</sup> Socioeconomic status was defined as low if the highest reported parental education was “3-year high school general studies, junior college” or lower, and high if it was “regional technical college, 4-year university degree” or higher.

Information about psychiatric diagnoses (registered between 2008-2023) was retrieved from the Norwegian Patient Registry (NPR). We included the following diagnostic categories: F1 (disorders due to psychoactive substance use); F2 (schizophrenia, schizotypal and delusional disorders); F31 (bipolar affective disorder); F32-F34.1 (depressive disorders); F40-F43, F93.0, F93.1, F93.2 (anxiety and stress-related disorders); F50 (eating disorders); F90 (hyperkinetic disorders); F84 (pervasive developmental disorders) (see Supplementary Text 1 and Table S4 for details). The categories were chosen to reflect a broad range of disorders relevant to adolescent and young adult mental health, and for which publicly available, well-powered GWAS data exist. Thus, our study included participants with at least one of the selected diagnoses, or no psychiatric diagnoses registered in the NPR. Individuals with psychiatric diagnoses outside the selected categories, but none within them, were excluded from the analysis.

Self-reported symptoms of mental health problems were assessed within the group of participants without any registered psychiatric diagnoses, as well as among participants with at least one diagnosis registered. The following instruments were used: Short Mood and Feeling Questionnaire (SMFQ),^15^ Screen for Child Anxiety Related Disorders (SCARED),^16^ and Eating Disorder Examination Questionnaire (EDE-Q).^17^ For behavioural assessment we used the ADHD-related items of the Parent/Teacher Rating Scale for Disruptive Behaviour Disorders (RS-DBD),^18^ and the prosocial subscale of the Strengths and Difficulties Questionnaire (SDQ),^19^ filled by mothers. Items were reverse-coded where necessary so that high scores reflected greater symptom load. Detailed information about the instruments is presented in the supplement (Table S2).

### Statistical Analysis

We used logistic regression to examine associations between screen time (explanatory variable) and presence of a psychiatric diagnosis (outcome variable). This was first done across all diagnostic categories combined, then separately for each individual diagnostic category with a prevalence greater than 1% in the study sample. We applied linear regression to assess the relationship between screen time and mental health symptom levels among participants without any psychiatric diagnoses. Age, sex assigned at birth, and SES were used as covariates in all regression models. Screen time use score was entered as a factor variable, with the category “1-2 h/day” set as a reference. Subsequently we conducted sex-stratified analysis by applying identical regression models to the female and male subsamples, with age and SES as covariates. Moreover, we conducted a sensitivity analysis restricting the outcome to diagnoses received one year or later after Q14-year completion, except for neurodevelopmental disorders.

To estimate PRS of psychiatric disorders we used summary statistics from recent large-scale GWASs of SCZ, BP, MD, ASD, ADHD, AUD, ANX and AN (Table S3). PRS were calculated using the reference-standardized, reproducible GenoPred pipeline.^20^ The sample for PRS analysis was restricted to unrelated participants of European ancestry.

We used multinomial logistic regression to examine whether participants with different levels of genetic liability to psychiatric disorders of interest, as reflected by PRS (explanatory variable), differed in their levels of screen-based activities (outcome variable). Within each model, age, genetic sex, and SES were included as covariates. All PRSs were pre-residualized for the first 10 genetic PCs and genotyping batch. To confirm that underlying associations were not driven by PRS of participants with psychiatric diagnoses, we performed a sensitivity PRS analysis for the subsample of individuals without a history of any mental disorder (see Supplementary Text 1 for details).

The degree of genetic confounding in the associations between moderate-to-high screen use and psychiatric diagnoses was estimated separately for females and males using a genetic sensitivity analysis called Gsens.^21^ This method fits Structural Equation Models based on the observed correlations between PRS, exposure and outcomes. Importantly, it allows adjustment of the correlation matrix under different scenarios, combining information on current PRS and single nucleotide polymorphism (SNP) or twin heritability estimates (Table S4). More details are provided in the supplement.

In all analyses, PRSs, adolescent-reported and mother-reported scores of mental health problems were standardized to mean 0 and SD 1. Original *p*-values for statistical significance are reported. We applied Bonferroni correction for multiple testing, adjusting the significance threshold based on the number of models tested within each analysis setting.

## Results

### Sample characteristics

Descriptive information on key study variables and basic demographic characteristics of the study sample is presented in Table 1. Detailed information about specific MoBa questions and variables is presented in Table S1, and Figures S2-S3.

Of 25,079 individuals with the Q-14year data and MBRN linkage, those with no psychiatric diagnoses within the selected categories, but at least one or more diagnoses outside them (*n* = 1289) were excluded. This resulted in a final analytic sample of 23,790 individuals (see Figure S1 for a study sample flow diagram), of which 3829 (16·09%) participants had at least one psychiatric diagnosis within the selected categories registered in the NPR. Further breakdowns by diagnostic subgroup, as well as information stratified by sex assigned at birth, is presented in Tables S5-S8.

### Association between mental illness and time spent on screen-based activities

Likelihood ratio test comparing the full models (including screen use variable) to the baseline models (including only age, sex assigned at birth, and SES) indicated significant associations for TV watching, gaming, and social media use (Figure 1, Table S9). TV watching for 3-4 h/day or more was linked to higher odds of having any psychiatric diagnosis, peaking at OR 2·36 (95% CI: 1·79, 3·10) for the maximum time spent. For gaming, a graded association was observed: adolescents with the lowest gaming levels had reduced odds of having a psychiatric diagnosis (OR 0·74, 95% CI: 0·64, 0·80), while those gaming 3-4 h/day or more showed increased odds, reaching OR 3·16 (95% CI: 2·55, 3·91) at the highest level. For social media use, we observed a U-shaped pattern: adolescents with both the least and the most time spent had higher odds of having a psychiatric diagnosis: OR 3·26 (95% CI: 2·73, 3·90) and 1·85 (95% CI: 1·59, 2·16), respectively. Sex-stratified analyses yielded generally consistent results (Figure S4, Table S10), except a different pattern of associations for gaming. For females, least time spent on gaming was linked to lower odds of having a psychiatric diagnosis (OR 0·62, 95% CI: 0·54, 0·71), whereas for males it was associated with slightly elevated odds (OR 1·44, 95% CI: 1·08, 1·91). A sensitivity analysis restricted to diagnoses received one year or later after Q14-year completion (excluding neurodevelopmental disorders) yielded consistent results, with associations between screen use and psychiatric diagnoses closely following those observed in the full analysis (Table S11, Figure S5).

**Figure 1.**
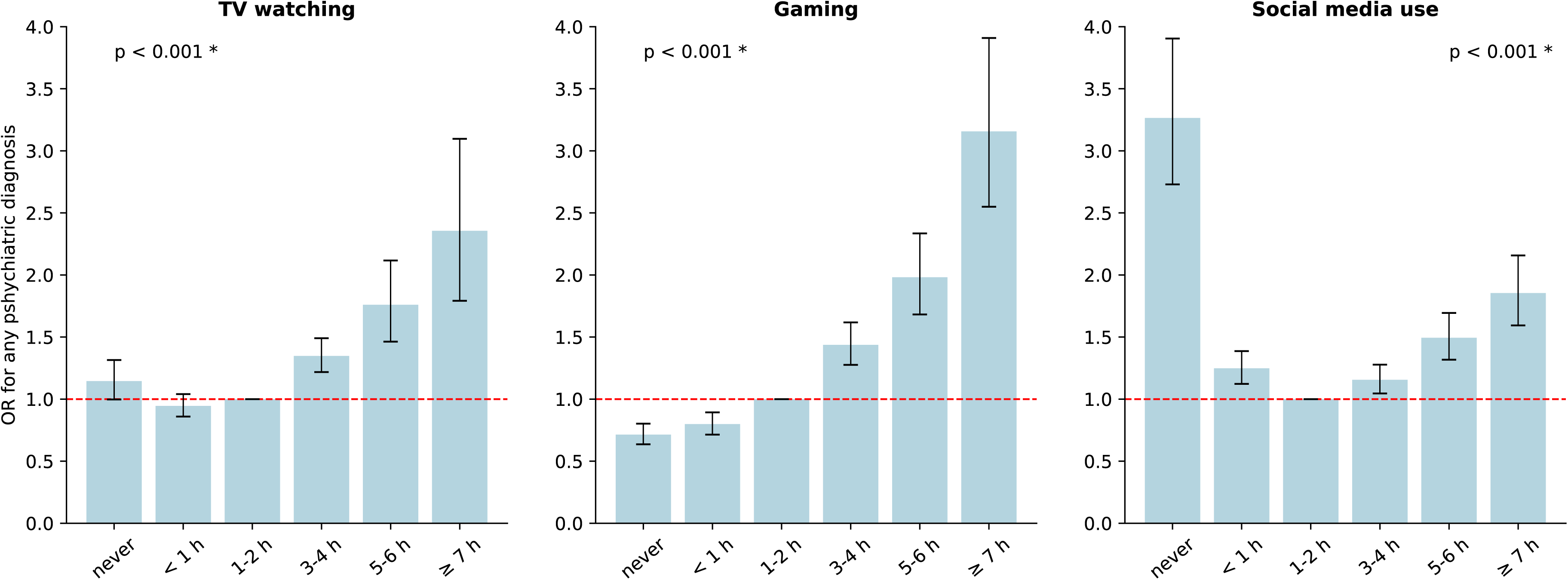
Associations between levels of screen-based activities and lifetime psychiatric diagnoses (*n* = 21,598). Odds ratios of having any psychiatric diagnosis (within the selected diagnostic categories) among participants with different levels of screen-based activities were estimated using logistic regression models with age, sex assigned at birth, and socioeconomic status as covariates. The “1–2 hours per day” group was used as a reference. Analyses were restricted for participants without shared mother ID. P-values from the likelihood ratio test comparing models with and without screen use variable are reported above each panel, with asterisks indicating statistically significant results after comparison to the family-wise error rate (α = 0·017), adjusted using Bonferroni correction (n_tests_ = 3). The horizontal line indicates no association (OR = 1). Error bars represent 95% confidence intervals.

Figure 2 and Table S12 present results across the diagnostic categories. Social media use was associated with elevated odds of having a diagnosis across all categories, while TV watching and gaming were significant predictors for all categories except eating disorders. Low gaming time was associated with slightly reduced odds across most diagnostic groups. Gaming for 3-4 h/day or more was linked to higher odds of having a diagnosis across all categories except eating disorders, with ORs ranging from 3·31 (95% CI: 2·46, 4·46) to 6·56 (95% CI: 4·54, 9·50) in the ≥ 7 h/day group. A similar pattern was observed for high social media use, although no association was found with pervasive developmental disorders. Notably, minimal social media use was associated with elevated odds across all categories, including eating disorders (OR 2·94, 95% CI: 1·80, 4·81) and pervasive developmental disorders (OR 10·68, 95% CI: 7·89, 14·45). Sex-stratified analyses (Table S13) showed that associations between screen behaviours—particularly gaming and social media use—and the odds of having a psychiatric diagnosis across most categories remained significant, largely following the pattern observed in the full sample (Figure S6).

**Figure 2.**
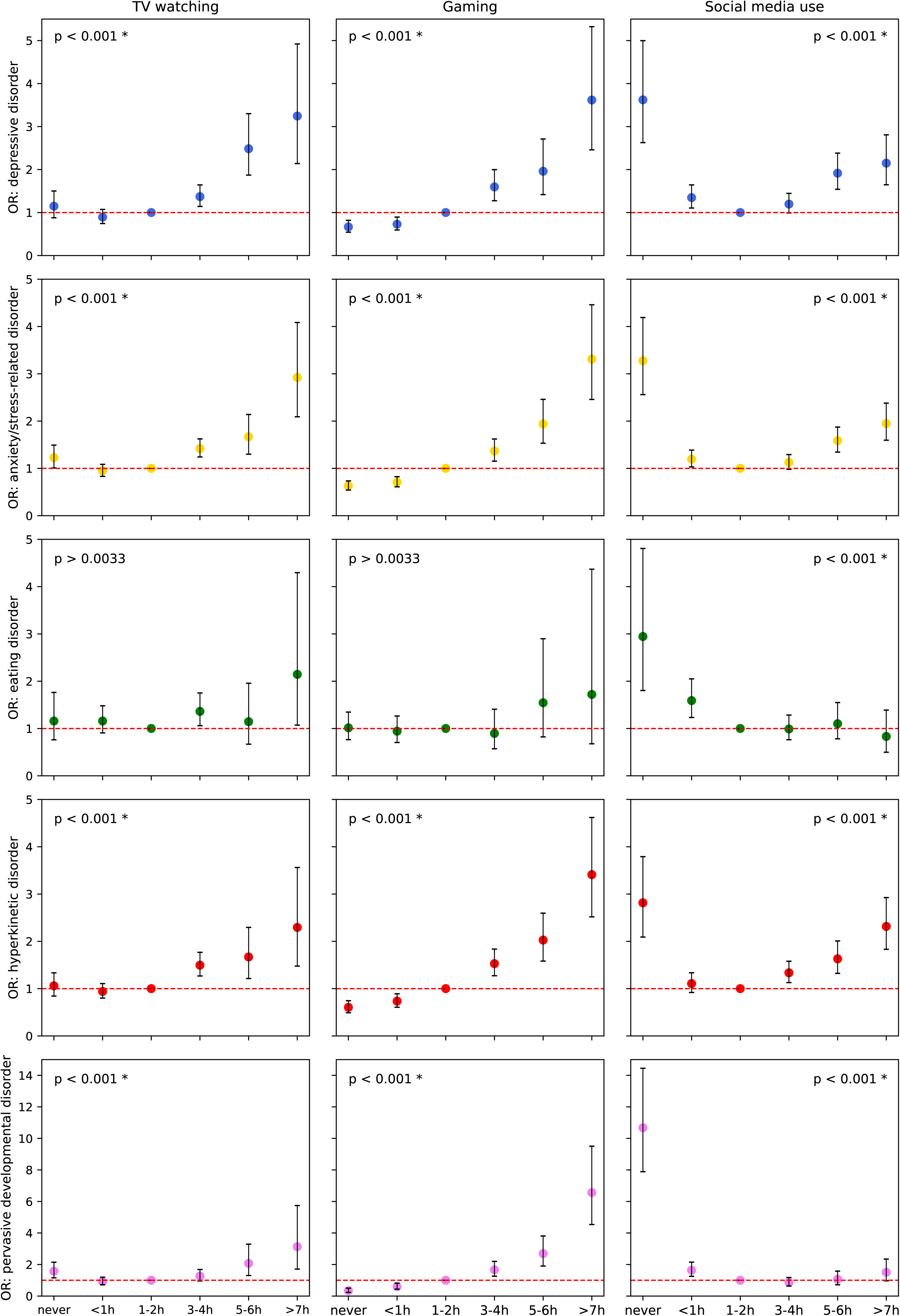
Associations between levels of screen-based activities and lifetime psychiatric diagnoses, stratified by a diagnostic category (*n* = 21,598). Odds ratios of having a psychiatric diagnosis (within specific diagnostic category) among participants with different levels of screen-based activities were estimated using logistic regression models with age, sex assigned at birth, and socioeconomic status as covariates. The “1–2 hours per day” group was used as a reference. Analyses were performed only for diagnostic categories with a prevalence greater than 1% in the study sample. P-values from the likelihood ratio tests comparing models with and without screen use variable are reported above each panel, with asterisks indicating statistically significant results after comparison to the family-wise error rate (α = 0·0033), adjusted using Bonferroni correction (n_tests_ = 15). The horizontal line indicates no association (OR = 1). Error bars represent 95% confidence intervals.

We also examined associations between screen use and self-reported mental health symptom scores in adolescents without any psychiatric diagnoses (Figure 3). All three screen behaviors were significantly associated with symptom levels (Table S14), consistent with the observations for clinical diagnoses. While effect sizes were modest, those spending 3–4 h/day or more on screens consistently reported higher symptom scores. Specifically, for associations between the highest social media use and depressive, anxiety, eating, and behavioural problem symptoms, the standardized regression coefficients (β) were estimated as β = 0·38 (95% CI: 0·32, 0·44), β = 0·25 (95% CI: 0·19, 0·31), β = 0·29 (95% CI: 0·23, 0·35), and β = 0·26 (95% CI: 0·20, 0·31), respectively, indicating differences in standardized symptom scores compared to the reference group. While screen use was not linked to diagnoses of eating disorders, all screen behaviours were positively associated with self-reported eating disorder symptoms (p < 0·001). Low gaming time was generally associated with lower symptom levels, with β = −0·17 (95% CI: −0·21, −0·13) for depressive symptoms. Conversely, the lowest level of social media use was linked to slightly elevated depression and anxiety scores: β = 0·13 (95% CI: 0·05, 0·21) for both, following the non-linear pattern seen in clinical outcomes. However, this pattern was not reflected in behaviour problem scores: although the lowest social media use group had higher odds of a hyperkinetic disorder diagnosis, no corresponding association was observed for self-reported behavioural symptoms. Overall, the patterns of association were consistent across symptom domains for each type of screen activity and generally aligned with findings for clinical diagnoses. Sex-stratified analyses retained many of these associations, particularly among females (Figure S7, Table S15). Symptom patterns among adolescents with psychiatric diagnoses also resembled those in the healthy subsample (Figure S8, Table S16).

**Figure 3.**
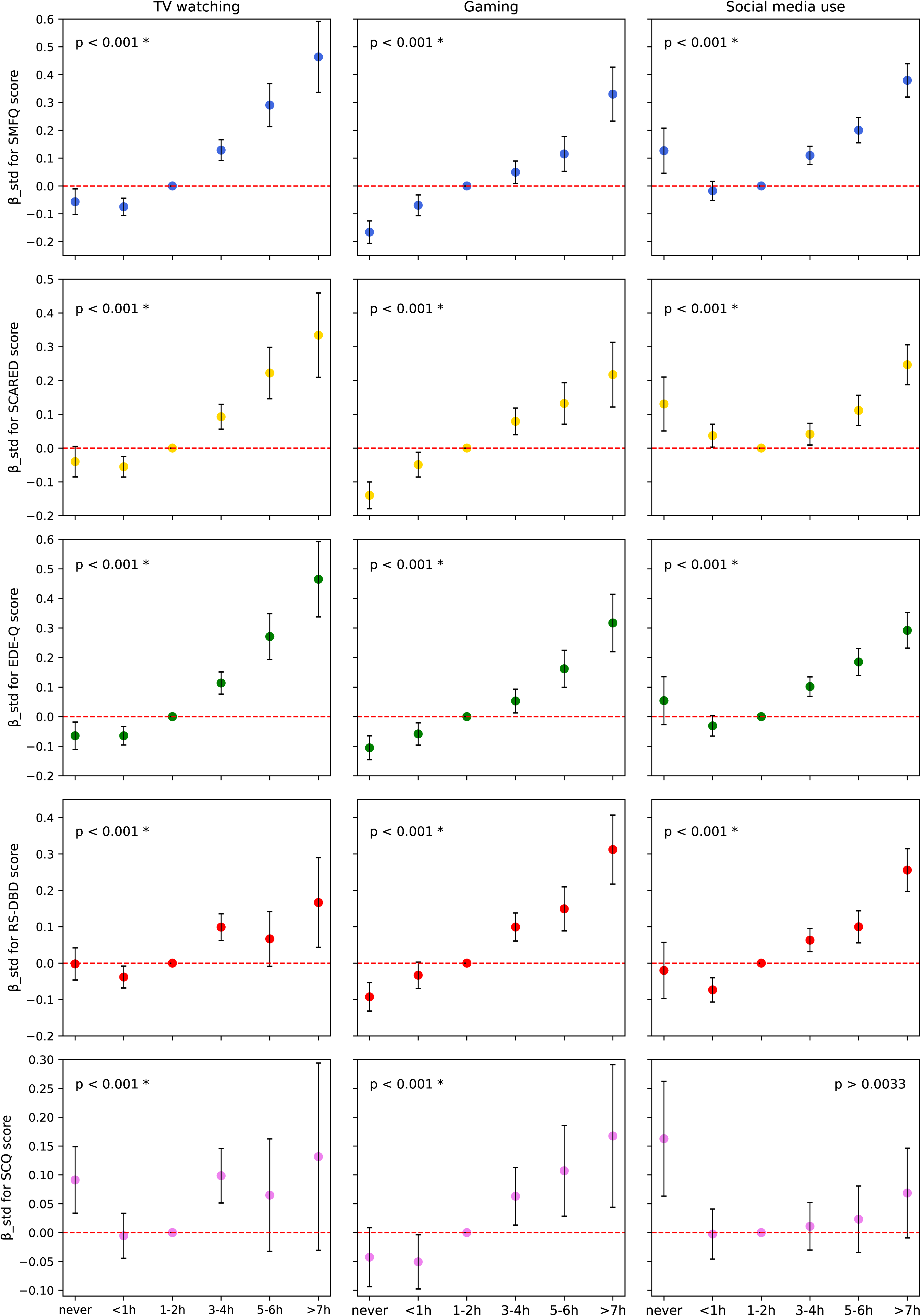
Associations between levels of screen-based activities and standardized symptom scores among participants without psychiatric diagnoses (n = 18,109). Associations between levels of screen-based activities and standardized symptom scores were estimated using linear regression models with age, sex assigned at birth, and socioeconomic status as covariates. The “1–2 hours per day” group was used as a reference. P-values from F-tests comparing models with and without screen use variable are reported above each panel, with asterisks indicating statistically significant results after comparison to the family-wise error rate (α = 0·0033), adjusted using Bonferroni correction (n_tests_ = 15). The horizontal line indicates no difference from the reference group (β = 0). Error bars represent 95% confidence intervals.

### Association between time spent on screen-based activities and PRS for psychiatric disorders

We identified several statistically significant associations between screen behaviors and PRS for major psychiatric disorders (Figure 4, Table S17). PRS_MD_ and PRS_ADHD_ were associated with all screen behaviors, while PRS_ASD_ and PRS_AN_ were associated only with gaming and social media use. One standard deviation (SD) increase in the PRS_MDD_ and PRS_ADHD_ was associated with a slightly higher odds of reporting the highest social media use level (OR 1·17, 95% CI: 1·09, 1·25, and OR 1·19, 95% CI: 1·11, 1·28, respectively), compared to reporting the reference level of screen use (“1-2 h/day”). Lowest level of social media use was associated with increased PRS_ASD_: one SD increase was linked to a higher likelihood of reporting no social media use (OR 1·24, 95% CI: 1·11, 1·28). Overall, the association patterns for PRS_MDD,_ PRS_ADHD_, and PRS_ASD_ with gaming and social media use reflected those observed for both clinical diagnoses and self-reported symptoms. By contrast, the pattern of associations for PRS_AN_ was opposite to that seen for eating disorder symptoms: a one SD increase in PRS_AN_ was associated with lower odds of reporting high engagement in gaming and social media (OR 0·89, 95% CI: 0·83, 0·96), and slightly higher odds of reporting rare or no gaming (OR 1·10, 95% CI: 1·05, 1·15). We have also observed associations between screen use and PRS_SCZ_, PRS_BP_, and PRS_AUD_, despite low clinical prevalence of the respective diagnoses in the analytical sample (Figure S9, Table S18). Sensitivity analysis performed for participants without a history of any psychiatric disorder produced concordant results (Figure S10, Tables S19).

**Figure 4.**
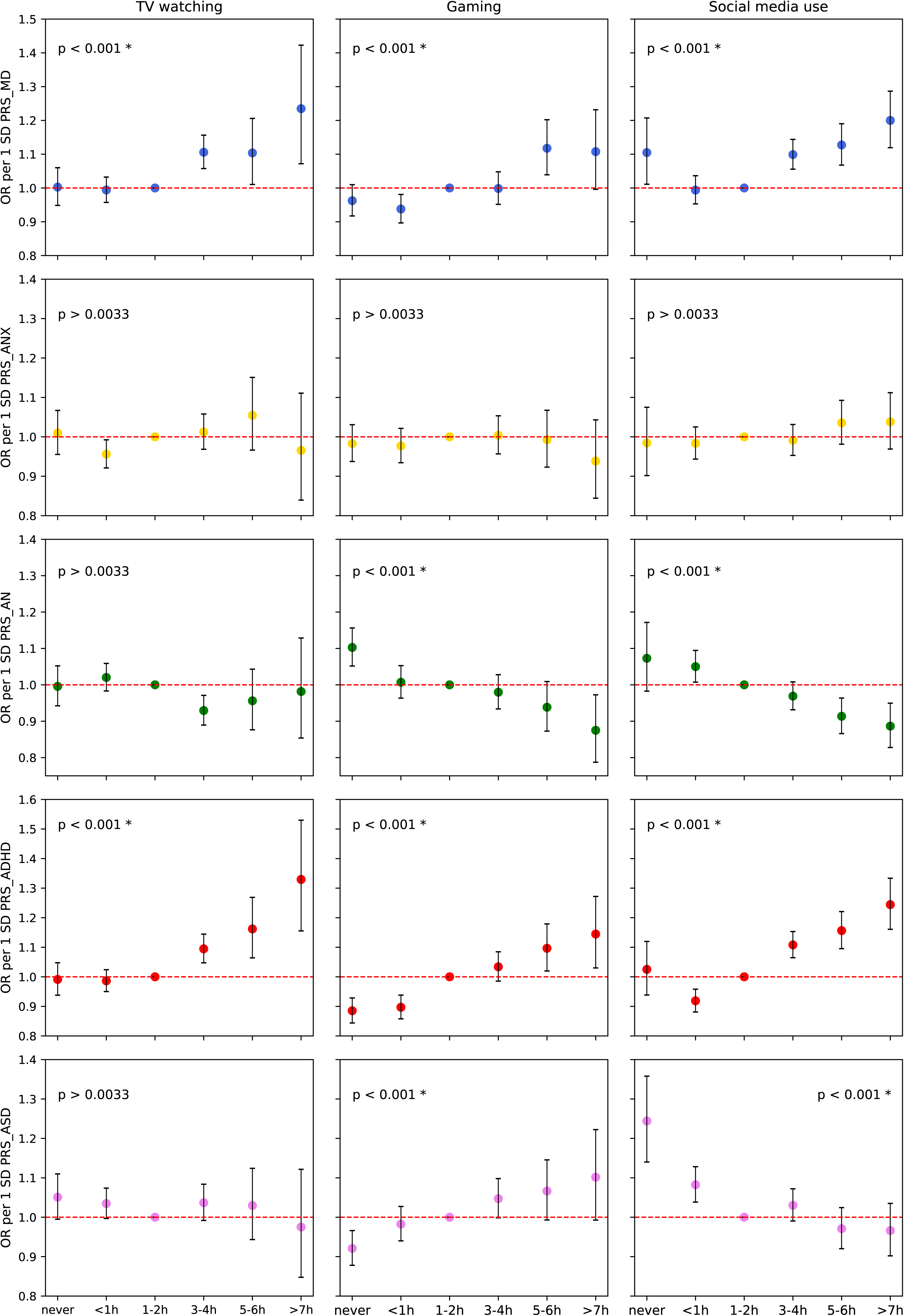
Associations between levels of screen-based activities and polygenic risk scores for major psychiatric disorders (*n* = 17,214). Associations between levels of screen-based activities and polygenic risk scores for major psychiatric disorders were estimated using multinomial regression models, with screen use modeled as a categorical outcome variable with six levels (reference category set at “1–2 hours per day”). All models were adjusted for the first 10 genetic PCs and genotyping batch, and within each model age, genetic sex, and socioeconomic status were included as covariates. Analyses were performed only for diagnostic categories with a prevalence greater than 1% in the study sample. P-values from the likelihood ratio tests comparing models with and without polygenic risk score are reported above each panel, with asterisks indicating statistically significant results after comparison to the family-wise error rate (α = 0·0033), adjusted using Bonferroni correction (n_tests_ = 15). The horizontal line indicates no association (OR = 1). Error bars represent 95% confidence intervals. MD: major depression; ANX: anxiety disorder; AN: anorexia nervosa; ADHD: attention-deficit hyperactivity disorder; ASD: autism spectrum disorder; PRS: polygenic risk score

### Genetic sensitivity analysis

Using Gsens models adjusted for SNP-based heritability, we found that a substantial proportion of the observed associations between psychiatric diagnoses and screen behaviours at the moderate-to-high level (i.e. 1-2 h/day or more) can likely be attributable to genetic factors—for example, up to 63·3% for social media use and hyperkinetic disorder diagnoses among females—though the phenotypic associations still remained statistically significant after accounting for this (Tables S20-S25, Figures S11-S16). When using twin-based heritability estimates, the extent of genetic confounding was large enough to fully attenuate the observed associations. Similar patterns of genetic confounding were observed across all evaluated screen behaviours and diagnoses, including depressive disorders, hyperkinetic disorders, and pervasive developmental disorders.

## Discussion

In this study, we leveraged data from a large, genotyped population-based cohort, and found that adolescent screen use is significantly associated with both psychiatric diagnoses and mental health traits. The associations varied by type of screen activity, the mental health phenotype, and the level of screen use. In addition, the associations were partly attributable to common genetic variants associated with psychiatric disorders, suggesting that shared genetic susceptibility may contribute to the observed associations between screen use and mental health phenotypes.

Across all screen-based activities, spending 3–4 hours or more per day was consistently associated with increased odds of psychiatric diagnoses—particularly depressive, anxiety/stress-related, and hyperkinetic disorders. A similar association was found with higher mental symptom scores among adolescents without a psychiatric diagnosis, suggesting a continuum of subclinical behaviours to diagnoses. While previous studies have reported associations between screen use and internalizing and externalizing symptoms,^22,23^ as well as autism spectrum disorder, ^24^ we provide a more comprehensive analysis across multiple mental health phenotypes, ranging from mental traits to clinical diagnoses. The concordant patterns of association across clinical diagnoses and subclinical mental health traits underscore the robustness of the findings.

A potential explanation of our findings may be that adolescents with pre-existing mental health problems exhibit different patterns of screen use, raising the possibility of reverse causation. Indeed, adolescents with internalizing and externalizing mental health conditions have been shown to spend more time on social media and engage with these platforms in qualitatively different ways.^25^ However, in our study the association between internalizing disorder diagnoses and social media use remained significant also when restricting analyses to diagnoses received one year or later after the questionnaire completion. Moreover, two small randomized controlled trials have shown that reducing screen use can improve depressive and internalizing symptoms in youth.^26,27^ This suggests that the observed associations are not solely attributable to pre-existing mental health conditions.

Our study revealed compelling associations between mental traits and disorders and very low screen use – an area often overlooked as most findings relate to excessive screen time. The lowest level of social media engagement was associated with increased odds of all psychiatric diagnoses, a pattern that was also reflected in higher self-reported internalizing symptoms and lower parent-reported prosocial behaviour. The association between low social media use and pervasive developmental disorders was particularly strong and has also been mentioned in a recent systematic review.^24^ Our findings suggest that minimal social media use may result from difficulties in peer engagement, reflecting impaired social functioning—a core feature of many mental disorders. The issue may be not low screen time itself, but rather difficulties in using digital platforms in socially rewarding ways – as suggested by a recent qualitative study involving autistic adolescents.^28^ Our findings support the notion that moderate restrictions on social media, when combined with targeted support and guidance, may help alleviate peer-related pressure to use social media platforms and promote healthier digital and in-person socialization.

We found that less time spent on gaming was linked to reduced odds of having a psychiatric diagnosis in females but not in males, which was also reflected in self-reported mental health traits. One possible explanation is that for adolescent girls, lower gaming may reflect engagement in a broader range of social or offline activities associated with better mental health, whereas in boys, gaming may be more socially normative and not necessarily linked to a psychopathology risk. While most other associations in our study showed similar patterns across females and males—despite differences in their overall screen use—the findings for gaming highlight the importance of considering sex-specific nuances in screen use research and caution against universal interpretations of screen time impacts across genders.

Our phenotypic and genetic findings showed overall alignment, suggesting genetic factors being involved. Some discrepancies emerged, as we found that higher genetic liability for depression and ADHD (PRS_MD_ and PRS_ADHD_), was associated with increased screen use, whereas no such link was observed for anxiety (PRS_ANX_). This may indicate that anxiety-related screen behaviours are more environmentally driven. We also found that higher genetic liability for anorexia nervosa (PRS_AN_) was associated with lower levels of both gaming and social media engagement. While this contrasts the positive phenotypic associations observed between self-reported eating disorder symptoms and screen use, it complies with the elevated odds of eating disorder diagnoses among individuals with minimal social media use. One possible explanation is that symptom-based measures may reflect a broader spectrum of eating behaviour difficulties, which are less severe than those required for a clinical diagnosis. Additionally, anorexia nervosa is often characterized by traits such as cognitive rigidity, high self-control, and social withdrawal—features that may contribute to disengagement from interactive, peer-oriented platforms like gaming and social media. It is also worth noting that the PRS_AN_ was derived from a GWAS with relatively low power, which may limit its interpretability.

Our study is not without limitations. Self-reported data are vulnerable to response bias,^29^ and it’s difficult to capture different dimensions of the screen-based activities such as active vs. passive engagement, the specific content consumed or social aspects of the activity. Ideally, future studies should integrate device- or platform-level trace data to provide a more comprehensive understanding of the relationships involved. Moreover, our study is mainly cross-sectional with 1 year follow up, and therefore cannot identify causal relationships between digital media use and mental health. However, even if reverse causation is present— particularly for neurodevelopmental disorders—this would not alter our findings regarding genetic confounding, and phenotypic associations would still be observed. Finally, selection bias is a challenge in cohort studies, which may limit the generalizability of findings.^30^

In conclusion, our findings point to a complex, bidirectional relationship between screen time and adolescent mental traits and disorders. We found distinct patterns of relationships across psychiatric diagnoses and types of screen use, and a U-formed relationship between level of screen time and mental disorders and traits. Our findings suggest shared genetics underlying the phenotypic associations between screen use and mental disorders, indicating genetic vulnerability to both. Additionally, the strong associations observed between minimal social media use and mental disorders may reflect impaired social functioning. Together, these findings can be used as basis for designing individualized approaches to screen use guidelines and preventive mental health interventions in youth.

## Acknowledgements

The Norwegian Mother, Father and Child Cohort Study is supported by the Norwegian Ministry of Health and Care Services and the Ministry of Education and Research. We are grateful to all the participating families in Norway who take part in this on-going cohort study.

For generating high-quality genomic data, we thank the Norwegian Institute of Public Health (NIPH), the HARVEST collaboration, the NORMENT Centre at the University of Oslo, the Center for Diabetes Research at the University of Bergen, deCODE Genetics, the Research Council of Norway, the SouthEastern and Western Norway Regional Health Authorities, the ERC AdG, Stiftelsen KG Jebsen, the Trond Mohn Foundation, and the Novo Nordisk Foundation

This work was performed on Services for sensitive data (TSD), University of Oslo, Norway, with resources provided by UNINETT Sigma2 - the National Infrastructure for High Performance Computing and Data Storage in Norway.

Disclaimer. Data from the Norwegian Patient Registry has been used in this publication. The interpretation and reporting of these data are the sole responsibility of the authors, and no endorsement by the Norwegian Patient Registry is intended nor should be inferred.

## Data Availability Statement

Data from the Norwegian Mother, Father and Child Cohort Study is managed by the Norwegian Institute of Public Health. Access requires approval from the Regional Committees for Medical and Health Research Ethics (REC), compliance with GDPR, and data owner approval. Participant consent does not allow individual-level data storage in repositories or journals. Researchers seeking access for replication must apply via www.helsedata.no.

## Ethical Consideration

All data and material in MoBa are collected with informed consent from participants in the study. Mothers participating in MoBa gave written informed consent on behalf of their children. At 18, the children are asked to give renewed consent for the continued use of data collected while they were younger. At 14, the adolescents were informed about the study and asked to provide new consent for participation in follow-up investigations. Data of participants who withdrew their consent before November 2024 were not included in the current study.

## Author’s Contributions

Concept and design (EF, OF, OAA, OBS); acquisition, analysis, or interpretation of data (all authors); methodology (EF, OF, NP, AAS, EH); software (OF, EH); supervision (HA, OAA, OBS); writing – original draft (EF, OBS), writing – review & editing (all authors).

