## Supplementary Note for "The phenotypic and genetic relationship between adolescent mental health and time spent on social media, gaming, and TV"

### Supplementary Text 1

*Phenotypic Variables*

Socioeconomic status (SES) was evaluated by extracting information about the highest level of parental education. At baseline, mothers were asked about the highest level of education they and their partners had completed (variables names AA1124 and AA1126, respectively, Q1 questionnaire). The maximum score (or the one available, in case one score was missing) was chosen to represent SES. For participants with both variables missing, we used data about the highest level of maternal education from the Q-8year questionnaire (variable name NN271).

SES was coded as 1 (“low’) if either the highest level of parental education in the Q1 questionnaire or maternal education in the Q-8year questionnaire was reported as score 4 (“3-year high school general studies, junior college”) or lower (see Table S1 for questions and variable names).

Information about adolescent mental health was retrieved both from the Norwegian Patient Registry (NPR), which contains ICD-10^1^ diagnoses registered in specialist health care services between 2008-2023, and from self-reports. Instruments used for assessing self-reported symptoms of mental health problems are presented in Table S2. A mean score of the items was computed for each instrument, requiring at least half the items to be non-missing and multiplied by the number of items in the instrument to give a representative score on the scale of the instrument. Items were reverse-coded where necessary so that high scores reflected greater symptom load.

*Statistical Analysis*

*Phenotypic Analysis*

For any pair of full siblings and maternal half-siblings (i. e. participants who had identical mother IDs), only one member was randomly retained in the analyses.

The results of the phenotypic analysis examining associations between screen time and lifetime psychiatric diagnoses are presented using odds ratios (OR) with 95% confidence intervals (CIs), with OR above 1.0 indicates increased odds of having a psychiatric diagnosis in a subgroup with a certain screen time use score, compared to a subgroup engaged in a particular screen-based activity for 1-2 hours per day.

When analysing each diagnostic category separately, individuals from the main analytical sample were included in the case group if they had at least one diagnosis within the category of interest, regardless of other diagnoses. The comparison group included only those with no psychiatric diagnoses.

*Polygenic risk score (PRS) analyses*

Full details about the genotyping and QC procedure are provided elsewhere.^2^ To estimate PRS of psychiatric disorders we used summary statistics from recent large-scale GWASs of schizophrenia (SCZ), bipolar disorder (BD), major depression (MD), autism spectrum disorder (ASD), attention-deficit hyperactivity disorder (ADHD), anxiety (ANX), anorexia nervosa (AN), and alcohol use disorder (AUD), independent of the MoBa sample^3–10^ (Table S3). For PRS calculation the sample was restricted to 17,216 unrelated participants of European ancestry following the GenoPred QC procedure.^11^ Subjects with ambiguous sex (based on genetics, *n* = 1) and subjects with chromosomal abnormalities (as indicated in the Medical Birth Registry of Norway, *n* = 1) were manually removed from analyses. Therefore, the final sample for PRS analysis was of 17,214 individuals (from 18,477 participants with both phenotypic and genetic data available). For association analyses we used PRSs calculated with SBayesR, one of the recommended methods for application in psychiatric disorders.^12^

The subsample for the sensitivity PRS analyses was restricted to individuals without a history of any psychiatric disorder, according to the NPR. Of the final PRS sample, 2,589 participants had at least one registered diagnosis, resulting in a sensitivity analysis sample of 14,625 individuals.

The results of the PRS analysis are presented using odds ratios (OR) with 95% CIs. As all PRSs were standardized to mean 0 and SD 1, OR above 1.0 indicates a higher odds of being in a given screen use category relative to the reference group, per one standard deviation increase in the respective PRS.

*Genetic sensitivity analysis*

Gsens framework was used to quantify genetic confounding for the associations between moderate-to-high screen use and psychiatric diagnoses. Screen use level was classified as moderate-to-high if the reported score was 3 or above (i.e. 1–2 hours per day or more). The number of participants with moderate-to-high use was 10,954 (6598 females, 4356 males) for TV watching, 8,451 (2204 females, 6247 males) for gaming, and 12,821 (7270 females, 5551 males) for social media use.

The associations of interest were adjusted on the polygenic scores for the outcome (GsensY). Structural equation models were fitted under three scenarios. In the first, the unadjusted polygenic score for the outcome was used. The second scenario assumed that the polygenic score captured the SNP-heritability of the outcome, whereas the third assumed that it accounted for the twin heritability representing the upper bound of potential genetic confounding. Heritability estimates used in Gsens analyses are presented in Table S4. Analyses were performed separately for females and males. All PRSs were pre-residualized for the first 10 genetic PCs and genotyping batch.

Correlations between polygenic scores, exposures, and outcomes were estimated using the lavCor function from the *lavaan* package in R. Ordinal and binary variables were specified using the “ordered” argument to apply appropriate correlation types. Gsens models were fitted only when all pairwise correlations between polygenic score, exposure, and outcome exceeded 0.03, to ensure model identifiability and stability. The correlation matrices are shown in Figures S10-S12.

Analyses were done using Python ver. 3.9.5, and R ver. 4.1.1.

### Table S1. Specific MoBa questions used in the study and variable names.

| **Question** | **Response options** | **Variable name** | **Questionnaire** |
| --- | --- | --- | --- |
| How much time do you usually spend during one weekday on the following activities? | | | |
| 1. Watch movies/series/TV | **1** - Never/ rarely  **2** - Less than 1 hour  **3** - 1-2 hours  **4** - 3-4 hours  **5** - 5-6 hours  **6** - 7 hours or more | UB18 | Q-14year |
| 2. Playing games (on PC, TV, tablet, mobile etc.) | **1** - Never/ rarely  **2** - Less than 1 hour  **3** - 1-2 hours  **4** - 3-4 hours  **5** - 5-6 hours  **6** - 7 hours or more | UB20 | Q-14year |
| 3. Sitting/lying down with PC, mobile or tablet (irrespective of activity) | **1** - Never/ rarely  **2** - Less than 1 hour  **3** - 1-2 hours  **4** - 3-4 hours  **5** - 5-6 hours  **6** - 7 hours or more | UB21 | Q-14year |
| 4. Communicating with friends on social media | **1** - Never/ rarely  **2** - Less than 1 hour  **3** - 1-2 hours  **4** - 3-4 hours  **5** - 5-6 hours  **6** - 7 hours or more | UB22 | Q-14year |
| Which of the following alternatives best describe your current gender identity? | Girl | UB232 | Q-14year |
|  | Boy | UB233 | Q-14year |
|  | Trans person | UB234 | Q-14year |
|  | Don’t know | UB235 | Q-14year |
|  | Do not wish to answer | UB236 | Q-14year |
| What education do you and the baby’s father have? (Fill in the highest level of education you have completed.) | | | |
| Education you have completed | **1** - 9-year secondary school  **2** - 1-2 year high school  **3** - Vocational high school  **4** - 3-year high school general studies, junior college  **5** - Regional technical college, 4-year university degree (bachelor’s degree, nurse, teacher, engineer)  **6** - University, technical college, more than 4 years (master’s degree, medical doctor, PhD) | AA1124 | Q1 |
| Education baby’s father has completed | **1** - 9-year secondary school  **2** - 1-2 year high school  **3** - Vocational high school  **4** - 3-year high school general studies, junior college  **5** - Regional technical college, 4-year university degree (bachelor’s degree, nurse, teacher, engineer)  **6** - University, technical college, more than 4 years (master’s degree, medical doctor, PhD) | AA126 | Q1 |
| What is your highest level of completed education? | **1** - 9-year secondary school  **2** - 1-2 years in high school  **3** - Vocational high school  **4** – General studies, 3-year high school  **5** – College, university up to 4 years  **6** – College, university more than 4 years | NN271 | Q-8year |

### Table S2. Instruments for assessing self-reported symptoms of mental health problems.

| **Question** | **Response options** | **Variable name** | **Questionnaire** |
| --- | --- | --- | --- |
| **Short Mood and Feelings Questionnaire (SMFQ)** | | | |
| Here follows a list of different disturbing feelings and thoughts one might have sometimes. Think about the past two weeks and mark each item whether you have felt or thought these ways. | | | |
| 1. Felt miserable or unhappy | **1**-Not true  **2**- Sometimes true  **3**-True | UB41 | Q-14year |
| 2. Felt so tired that I just sat around and did nothing |  | UB42 |  |
| 3. Was very restless |  | UB43 |  |
| 4. Didn’t enjoy anything at all |  | UB44 |  |
| 5. Felt I was no good anymore |  | UB45 |  |
| 6. Cried a lot |  | UB46 |  |
| 7. Hated myself |  | UB47 |  |
| 8. Thought I could never be as good as other kids |  | UB48 |  |
| 9. Felt lonely |  | UB49 |  |
| 10. Thought nobody really loved me |  | UB50 |  |
| 11. Felt I was a bad person |  | UB51 |  |
| 12. Felt I did everything wrong |  | UB52 |  |
| 13. Found it hard to think/concentrate |  | UB53 |  |
| **Screen for Child Anxiety Related Disorders (SCARED)** | | | |
| Children and youth might be anxious at times or be bothered by strange thoughts. Consider the past months and mark each item the way that best applies to you. | | | |
| 1. I have been really frightened for no reason at all | **1**-Not true  **2**- Sometimes true  **3**-True | UB166 | Q-14year |
| 2. I have been afraid to be alone in the house |  | UB167 |  |
| 3. People have told me that I worry too much |  | UB168 |  |
| 4. I have been scared to go to school |  | UB169 |  |
| 5. I have been shy |  | UB170 |  |
| **Selected questions from the** **Eating Disorder Examination Questionnaire (EDE-Q)** | | | |
| Respond to each question: | | | |
| 1. When you think about the past 4 weeks, how often have you been deliberately trying to limit the amount of food you eat to influence your shape or weight? | **1**-Never/rarely  **2**- Sometimes  **3**-Often  **4**-Very often | UB79 | Q-14year |
| 2. Over the past 4 weeks, how often have you tried to follow definite rules regarding what you can eat, in order to influence your shape or weight (for example a limited amount of calories)? |  | UB80 |  |
| 3. Over the past 4 weeks, how often have you had a definite fear of losing control over eating? |  | UB81 |  |
| 4. Over the past 4 weeks, has thinking about food, eating or calories made it very difficult to concentrate on things you are interested in (for example, working, following a conversation, or reading)? |  | UB82 |  |
| 5. Over the past 4 weeks, have you eaten secretly? |  | UB83 |  |
| 6. How dissatisfied have you been with your shape (what you see in the mirror)? |  | UB84 |  |
| 7. How uncomfortable have you felt seeing your own body (for example seeing your shape in the mirror, while undressing, taking a bath or shower? |  | UB85 |  |
| 8. How uncomfortable have you felt about others seeing your shape or figure (for example in communal changing rooms, when swimming or wearing tight clothes)? |  | UB86 |  |
| **Parent/Teacher Rating Scale for Disruptive Behaviour Disorders (RS-DBD)** | | | |
| Choose the alternative that best describes your child’s behaviour over the past 6 months | | | |
| 1. Fails to give close attention to details or makes careless mistakes in schoolwork | **1**-Never/rarely  **2**- Sometimes  **3**-Often  **4**-Very often | UM152 | Q-14year mother |
| 2. Has difficulty sustaining attention in tasks or play activities |  | UM153 |  |
| 3. Does not seem to listen when spoken to directly |  | Um154 |  |
| 4. Does not follow through on instructions and fails to finish school work, chores or duties (not due to oppositional behaviour or failure to understand instructions) |  | UM155 |  |
| 5. Has difficulty organizing tasks and activities |  | UM156 |  |
| 6. Avoids, dislikes or is reluctant to engage in tasks that require sustained mental effort (such as schoolwork or homework) |  | UM157 |  |
| 7. Loses things necessary for tasks or activities (pencils, books, toys) |  | UM158 |  |
| 8. Is easily distracted |  | UM159 |  |
| 9. Is forgetful in daily activities |  | UM160 |  |
| 10. Fidgets with hands or feet or squirms in seat (sits uneasily) |  | UM161 |  |
| 11. Leaves seat in classroom or in other situations in which remaining seated is expected (e.g. at the table or in group gathering) |  | UM162 |  |
| 12. Runs about or climbs excessively in situations in which it is inappropriate |  | UM163 |  |
| 13. Has difficulty playing or engaging in leisure activities quietly |  | UM164 |  |
| 14. Is “on the go” or acts as if “driven by a motor” |  | UM165 |  |
| 15. Talks excessively |  | UM166 |  |
| 16. Blurts out answers before questions have been completed |  | UM167 |  |
| 17. Has difficulty awaiting turn |  | UM168 |  |
| 18. Interrupts or intrudes on others, such as in conversation or play |  | UM169 |  |
| **Strengths and Difficulties Questionnaire (SDQ) – Prosocial Subscale** | | | |
| Give answers on the basis of your child’s behaviour over the past 6 months | | | |
| Is considerate to other people’s feelings | **1**-Not true  **2**- Somewhat true  **3**-Certainly true | UM189 | Q-14year mother |
| Shares readily with other youths (treats, games, other things) |  | UM190 |  |
| Is helpful if someone is hurt, upset or feeling ill |  | UM191 |  |
| Is kind to younger children/youths |  | UM192 |  |
| Often volunteers to help others (parents, teachers, other children/youths) |  | UM193 |  |

*The table provides a brief overview of the MoBa instruments used to assess symptoms of mental health problems in this study. Detailed information about the MoBa questionnaires, including descriptions of all instruments and variables, is available on the official website of the Norwegian Mother, Father, and Child Study:* [*https://www.fhi.no/op/studier/moba/*](https://www.fhi.no/op/studier/moba/)*.*

### Table S3. Overview of the GWASs contributing to the study

| **Phenotype** | **Abbreviation** | **Cases/controls** | **PubMedID** |
| --- | --- | --- | --- |
| Alcohol use disorder | AUD | 55,584/218,807 | 30940813 |
| Anorexia nervosa | AN | 16,992/55,525 | 31308545 |
| Anxiety disorders | ANX | 97,383/1,169,397 | 39294497 |
| Attention-deficit hyperactivity disorder | ADHD | 38,691/186,843 | 36702997 |
| Autism spectrum disorder | ASD | 18,381/27,969 | 30804558 |
| Bipolar disorder | BP | 41,917/371,549 | 34002096 |
| Major depression | MD | 412,305/1,588,397 | 34045744 |
| Schizophrenia | SCZ | 53,386/77,258 | 35396580 |

### Table S4. Heritability estimates used in genetic sensitivity analyses.

| **Phenotype** | **SNP heritability** | **PubMedID** | **Twin heritability** | **PubMedID** |
| --- | --- | --- | --- | --- |
| Major depression | 0.113 | 34045744 | 0.37 | 11007705 |
| Attention-deficit hyperactivity disorder | 0.14 | 36702997 | 0.74 | 29892054 |
| Autism spectrum disorder | 0.118 | 30804558 | 0.83 | 28973605 |

### Table S5. Percentage of psychiatric diagnoses for each main diagnostic subgroup in participants with Q-14year data and MBRN linkage (n = 25,079), with the maximum of three most common categories (for those not included in the study).

| **Diagnostic subgroup** | **N cases (%)** |
| --- | --- |
| Organic, including symptomatic, mental disorders (F00-F09) | <10 (<0.040%) |
| Other mental disorders due to brain damage and dysfunction and to physical disease (F06) | <10 (<0.040%) |
| Mental and behavioural disorders due to psychoactive substance use (F10-F19) | 215 (0.86%) |
| Schizophrenia, schizotypal and delusional disorders (F20-F29) | 55 (0.22%) |
| Mood disorders (F30-F39) | 1042 (4.16%) |
| Neurotic, stress-related and somatoform disorders (F40-F49) | 2055 (8.19%) |
| Behavioral syndromes associated with physiological disturbances and physical factors (F50-F59) | 484 (1.93%) |
| Disorders of adult personality and behaviour (F60-F69) | 77 (0.31%) |
| Specific personality disorders (F60) | 77 (0.31%) |
| Mental retardation (F70-F79) | 48 (0.19%) |
| Mild mental retardation (F70) | 34 (0.14%) |
| Moderate mental retardation (F71) | 13 (0.052%) |
| Unspecified mental retardation (F79) | 10 (0.040%) |
| Disorders of psychological development (F80-F89) | 946 (3.77%) |
| Specific developmental disorders of speech and language (F80) | 179 (0.71%) |
| Specific developmental disorders of scholastic skills (F81) | 331 (1.32%) |
| Pervasive developmental disorders (F84) | 472 (1.88%) |
| Behavioral and emotional disorders with onset usually occurring in childhood and adolescence (F90-F98) | 2597 (10.36%) |
| Hyperkinetic disorders (F90) | 1210 (4.82%) |
| Emotional disorders with onset specific to childhood (F93) | 424 (1.69%) |
| Other behavioral and emotional disorders with onset usually occurring in childhood and adolescence (F98) | 864 (3.45%) |

### Table S6. Percentage of psychiatric diagnoses for each main diagnostic subgroup in participants with Q-14year data and MBRN linkage (*n* = 25,079), stratified by sex assigned at birth.

| **Diagnostic subgroup** | **N cases (%), females** | **N cases (%), males** |
| --- | --- | --- |
| Organic, including symptomatic, mental disorders (F00-F09) | <10 (<0.074%) | <10 (<0.086%) |
| Mental and behavioural disorders due to psychoactive substance use (F10-F19) | 108 (0.80%) | 107 (0.92%) |
| Schizophrenia, schizotypal and delusional disorders (F20-F29) | 31 (0.23%) | 24 (0.21%) |
| Mood disorders (F30-F39) | 781 (5.82%) | 261 (2.24%) |
| Neurotic, stress-related and somatoform disorders (F40-F49) | 1538 (11.46%) | 517 (4.44%) |
| Behavioral syndromes associated with physiological disturbances and physical factors (F50-F59) | 451 (3.36%) | 33 (0.28%) |
| Disorders of adult personality and behaviour (F60-F69) | 73 (0.54%) | <10 (0.034%) |
| Mental retardation (F70-F79) | 24 (0.18%) | 24 (0.21%) |
| Disorders of psychological development (F80-F89) | 409 (3.05%) | 537 (4.61%) |
| Pervasive developmental disorders (F84) | 201 (1.50%) | 271 (2.33%) |
| Behavioral and emotional disorders with onset usually occurring in childhood and adolescence (F90-F98) | 1272 (9.47%) | 1325 (11.37%) |
| Hyperkinetic disorders (F90) | 610 (4.54%) | 600 (5.15%) |

### Table S7. Percentage of psychiatric diagnoses for each main diagnostic category selected for the current study in participants with Q-14year data and MBRN linkage, (*n* = 25,079).

| **Diagnostic subgroup** | **N cases (%)** |
| --- | --- |
| Mental and behavioural disorders due to psychoactive substance use (F10-F19) | 215 (0.86%) |
| Mental and behavioural disorders due to use of alcohol (F10) * | 146 (0.58%) |
| Mental and behavioural disorders due to use of opioids (F11) * | <10 (<0.040%) |
| Mental and behavioural disorders due to use of cannabinoids (F12) * | 45 (0.18%) |
| Mental and behavioural disorders due to use of sedatives or hypnotics (F13) * | <10 (<0.040%) |
| Mental and behavioural disorders due to use of cocaine (F14) * | <10 (<0.040%) |
| Mental and behavioural disorders due to use of other stimulants, including caffeine (F15) * | 15 (0.060%) |
| Mental and behavioural disorders due to use of hallucinogens (F16) * | <10 (<0.040%) |
| Mental and behavioural disorders due to use of tobacco (F17) * | <10 (<0.040%) |
| Mental and behavioural disorders due to multiple drug use and use of other psychoactive substances (F19) * | 33 (0.13%) |
| Schizophrenia, schizotypal and delusional disorders (F20-F29) | 55 (0.22%) |
| Schizophrenia (F20) * | <10 (<0.040%) |
| Schizotypal disorder (F21) * | <10 (<0.040%) |
| Persistent delusional disorders (F22) * | <10 (<0.040%) |
| Acute and transient psychotic disorders (F23) * | 11 (0.044%) |
| Schizoaffective disorders (F25) * | <10 (<0.040%) |
| Other nonorganic psychotic disorders (F28) * | <10 (<0.040%) |
| Unspecified nonorganic psychosis (F29) * | 29 (0.12%) |
| Mood disorders (F30-F39) | 1042 (4.16%) |
| Manic episode (F30) * | <10 (<0.040%) |
| Bipolar affective disorder (F31) * | 40 (0.16%) |
| Depressive episode (F32) * | 914 (3.64%) |
| Recurrent depressive disorder (F33) * | 126 (0.50%) |
| Persistent mood [affective] disorders (F34) | 44 (0.18%) |
| Dysthymia (F34.1) * | 40 (0.16%) |
| Other mood [affective] disorders (F38) | <10 (<0.040%) |
| Unspecified mood [affective] disorder (F39) | 12 (0.048%) |
| Neurotic, stress-related and somatoform disorders (F40-F49) | 2055 (8.19%) |
| Phobic anxiety disorders (F40) * | 685 (2.73%) |
| Other anxiety disorders (F41) * | 702 (2.80%) |
| Reaction to severe stress, and adjustment disorders (F43) * | 737 (2.94%) |
| Dissociative [conversion] disorders (F44) | 33 (0.13%) |
| Somatoform disorders (F45) | 87 (0.35%) |
| Other neurotic disorders (F48) | 41 (0.16%) |
| Behavioral syndromes associated with physiological disturbances and physical factors (F50-F59) | 484 (1.93%) |
| Eating disorders (F50) * | 484 (1.93%) |
| Disorders of psychological development (F80-F89) | 946 (3.77%) |
| Pervasive developmental disorders (F84) | 472 (1.88%) |
| Childhood autism (F84.0) * | 61 (0.24%) |
| Atypical autism (F84.1) * | 38 (0.15%) |
| Overactive disorder associated with mental retardation and stereotyped movements (F84.4) | <10 (<0.040%) |
| Asperger syndrome (F84.5) * | 328 (1.31%) |
| Other pervasive developmental disorders (F84.8) * | <10 (<0.040%) |
| Pervasive developmental disorder, unspecified (F84.9) * | 103 (0.41%) |
| Behavioral and emotional disorders with onset usually occurring in childhood and adolescence (F90-F98) | 2597 (10.36%) |
| Hyperkinetic disorders (F90) | 1210 (4.82%) |
| Disturbance of activity and attention (F90.0) * | 1137 (4.53%) |
| Hyperkinetic conduct disorder (F90.1) * | 67 (0.27%) |
| Other hyperkinetic disorders (F90.8) * | 76 (0.30%) |
| Hyperkinetic disorders, unspecified (F90.9) * | 52 (0.21%) |
| Emotional disorders with onset specific to childhood (F93) | 424 (1.69%) |
| Separation anxiety disorder of childhood (F93.0) * | 79 (0.32%) |
| Phobic anxiety disorder of childhood (F93.1) * | 26 (0.10%) |
| Social anxiety disorder of childhood (F93.2) * | 16 (0.063%) |

*Only categories with at least one registered case are shown. Asterisks indicate diagnostic categories included in the study analysis. Subcategory counts (e.g., F40-F45, F48) may exceed the total for the broader category (e.g., F4) because some participants received multiple diagnoses within the same diagnostic block. Each individual is counted only once at the broader category level.*

### Table S8. Percentage of psychiatric diagnoses in the final analytic sample (*n* = 23,790), shown for the full sample and separately by sex assigned at birth, across selected diagnostic categories.

| **Diagnostic subgroup** | **N cases (%)** | **N cases (%), females** | **N cases (%), males** |
| --- | --- | --- | --- |
| Mental and behavioural disorders due to psychoactive substance use (F10-F19) | 215 (0.90%) | 108 (0.84%) | 107 (0.98%) |
| Mental and behavioural disorders due to use of alcohol (F10) | 146 (0.61%) | 72 (0.56%) | 74 (0.68%) |
| Mental and behavioural disorders due to use of opioids (F11) | <10 (<0.042%) | <10 (<0.078%) | <10 (<0.092%) |
| Mental and behavioural disorders due to use of cannabinoids (F12) | 45 (0.19%) | 20 (0.16%) | 25 (0.23%) |
| Mental and behavioural disorders due to use of sedatives or hypnotics (F13) | <10 (<0.042%) | <10 (<0.078%) | <10 (<0.092%) |
| Mental and behavioural disorders due to use of cocaine (F14) | <10 (<0.042%) | <10 (<0.078%) | <10 (<0.092%) |
| Mental and behavioural disorders due to use of other stimulants, including caffeine (F15) | 15 (0.063%) | 10 (0.078%) | <10 (<0.092%) |
| Mental and behavioural disorders due to use of hallucinogens (F16) | <10 (<0.042%) | <10 (<0.078%) | <10 (<0.092%) |
| Mental and behavioural disorders due to use of tobacco (F17) | <10 (<0.042%) | <10 (<0.078%) | 0 (0.00%) |
| Mental and behavioural disorders due to multiple drug use and use of other psychoactive substances (F19) | 33 (0.14%) | 15 (0.12%) | 18 (0.16%) |
| Schizophrenia, schizotypal and delusional disorders (F20-F29) | 55 (0.23%) | 31 (0.24%) | 24 (0.21%) |
| Schizophrenia (F20) | <10 (<0.042%) | <10 (<0.078%) | <10 (<0.092%) |
| Schizotypal disorder (F21) | <10 (<0.042%) | <10 (<0.078%) | 0 (0.00%) |
| Persistent delusional disorders (F22) | <10 (<0.042%) | <10 (<0.078%) | <10 (<0.092%) |
| Acute and transient psychotic disorders (F23) | 11 (0.04%) | <10 (<0.078%) | <10 (<0.092%) |
| Schizoaffective disorders (F25) | <10 (<0.042%) | <10 (<0.078%) | 0 (0.00%) |
| Other nonorganic psychotic disorders (F28) | <10 (<0.042%) | <10 (<0.078%) | 0 (0.00%) |
| Unspecified nonorganic psychosis (F29) | 29 (0.12%) | 16 (0.12%) | 13 (0.12%) |
| Mood disorders (F30-F39) | 1033 (4.34%) | 774 (6.01%) | 259 (2.37%) |
| Manic episode (F30) | <10 (<0.042%) | <10 (<0.078%) | <10 (<0.092%) |
| Bipolar affective disorder (F31) | 40 (0.16%) | 32 (0.25%) | <10 (<0.092%) |
| Depressive episode (F32) | 914 (3.64%) | 684 (5.31%) | 230 (2.11%) |
| Recurrent depressive disorder (F33) | 126 (0.50%) | 101 (0.78%) | 25 (0.23%) |
| Persistent mood [affective] disorders (F34) | 43 (0.18%) | 33 (0.26%) | 10 (0.091%) |
| Dysthymia (F34.1) | 40 (0.16%) | 31 (0.24%) | <10 (<0.092%) |
| Neurotic, stress-related and somatoform disorders (F40-F49) | 1882 (7.91%) | 1423 (11.05%) | 459 (4.20%) |
| Phobic anxiety disorders (F40) | 685 (2.73%) | 535 (4.16%) | 150 (1.37%) |
| Other anxiety disorders (F41) | 702 (2.80%) | 557 (4.33%) | 145 (1.33%) |
| Reaction to severe stress, and adjustment disorders (F43) | 737 (2.94%) | 542 (4.21%) | 195 (1.79%) |
| Behavioral syndromes associated with physiological disturbances and physical factors (F50-F59) | 484 (2.03%) | 451 (3.50%) | 33 (0.30%) |
| Eating disorders (F50) | 484 (2.03%) | 451 (3.50%) | 33 (0.30%) |
| Disorders of psychological development (F80-F89) | 755 (3.17%) | 350 (2.72%) | 405 (3.71%) |
| Pervasive developmental disorders (F84) | 472 (1.88%) | 201 (1.56%) | 271 (2.48%) |
| Childhood autism (F84.0) | 61 (0.24%) | 12 (0.093%) | 49 (0.45%) |
| Atypical autism (F84.1) | 38 (0.15%) | 11 (0.085%) | 27 (0.25%) |
| Asperger syndrome (F84.5) | 328 (1.31%) | 154 (1.20%) | 174 (1.59%) |
| Other pervasive developmental disorders (F84.8) | <10 (<0.042%) | <10 (<0.078%) | <10 (<0.092%) |
| Pervasive developmental disorder, unspecified (F84.9) | 103 (0.41%) | 38 (0.30%) | 65 (0.60%) |
| Behavioral and emotional disorders with onset usually occurring in childhood and adolescence (F90-F98) | 1634 (6.87%) | 883 (6.86%) | 751 (6.88%) |
| Hyperkinetic disorders (F90) | 1210 (4.82%) | 610 (4.74%) | 600 (5.50%) |
| Disturbance of activity and attention (F90.0) | 1137 (4.53%) | 573 (4.45%) | 564 (5.17%) |
| Hyperkinetic conduct disorder (F90.1) | 67 (0.27%) | 24 (0.19%) | 43 (0.39%) |
| Other hyperkinetic disorders (F90.8) | 76 (0.30%) | 34 (0.26%) | 42 (0.38%) |
| Hyperkinetic disorders, unspecified (F90.9) | 52 (0.21%) | 24 (0.19%) | 28 (0.26%) |
| Emotional disorders with onset specific to childhood (F93) | 424 (1.69%) | 178 (1.38%) | 89 (0.82%) |
| Separation anxiety disorder of childhood (F93.0) | 79 (0.32%) | 52 (0.40%) | 27 (0.25%) |
| Phobic anxiety disorder of childhood (F93.1) | 26 (0.10%) | 18 (0.14%) | <10 (<0.092%) |
| Social anxiety disorder of childhood (F93.2) | 16 (0.063%) | 12 (0.09%) | <10 (<0.092%) |

*Subcategory counts may exceed the total for the broader category because some participants received multiple diagnoses within the same diagnostic block. Each individual is counted only once at the broader category level.*

### Table S9. Results of likelihood ratio tests comparing full models (including screen use score) to baseline models (including only age, sex assigned at birth, and socioeconomic status) (*n* = 21,598).

| **Phenotype** | **ΔDeviance** | ***p­*-value** |
| --- | --- | --- |
| TV watching | 101.33 | < 2.2e-16 * |
| Gaming | 275.35 | < 2.2e-16 * |
| Social media use | 202.25 | < 2.2e-16 * |

*Original p-values are reported, with asterisks indicating statistically significant results after comparison to the family-wise error rate (α = 0.017), adjusted using Bonferroni correction (n_tests_ = 3). Example of models compared: psychiatric_diagnosis ~ age + sex + socioeconomic_status vs. psychiatric_diagnosis ~ age + sex + socioeconomic_status + TV_watching_score.*

### Table S10. Results of likelihood ratio tests comparing full models (including screen use score) to baseline models (including only age and socioeconomic status), stratified by sex assigned at birth (*n* = 21,598).

| **Phenotype** | **Females (*n =* 11,742)** | | **Males (*n* = 9856)** | |
| --- | --- | --- | --- | --- |
|  | **ΔDeviance** | ***p­*-value** | **ΔDeviance** | ***p­*-value** |
| TV watching | 74.92 | 9.68e-15 * | 30.38 | 1.24e-05 * |
| Gaming | 172.26 | < 2.2e-16 * | 132.36 | < 2.2e-16 * |
| Social media use | 98.83 | < 2.2e-16 * | 98.83 | < 2.2e-16 * |

*Original p-values are reported, with asterisks indicating statistically significant results after comparison to the family-wise error rate (α = 0.0083), adjusted using Bonferroni correction (n_tests_ = 6). Example of models compared: psychiatric_diagnosis_females ~ age + socioeconomic_status vs. psychiatric_diagnosis_females ~ age + socioeconomic_status + TV_watching_score.*

### Table S11. Results of likelihood ratio tests comparing full models (including screen use score) to baseline models (including only age, sex assigned at birth, and socioeconomic status), restricted to diagnoses received one year or later after Q14-year completion (*n* = 18,976).

| **Phenotype** | **ΔDeviance** | ***p­*-value** |
| --- | --- | --- |
| TV watching | 19.95 | 0.0013 * |
| Gaming | 20.80 | 0.00089 * |
| Social media use | 27.17 | 5.29e-05 * |

*Diagnoses of neurodevelopmental disorders were excluded from analysis. Original p-values are reported, with asterisks indicating statistically significant results after comparison to the family-wise error rate (α = 0.017), adjusted using Bonferroni correction (n_tests_ = 3). Example of models compared: psychiatric_diagnosis_afterQ14completion ~ age + sex + socioeconomic_status vs. psychiatric_diagnosis_afterQ14completion ~ age + socioeconomic_status + sex + TV_watching_score.*

### Table S12. Results of likelihood ratio tests comparing full models (including screen use score) to baseline models (including only age, sex assigned at birth, and socioeconomic status), performed across specific diagnostic categories (*n* = 21,598).

| **Phenotype** | **ΔDeviance** | ***p­*-value** |
| --- | --- | --- |
| *Depressive disorders* | | |
| TV watching | 69.95 | 1.049e-13 * |
| Gaming | 107.92 | < 2.2e-16 * |
| Social media use | 86.972 | < 2.2e-16 * |
| *Anxiety/stress-related disorders* | | |
| TV watching | 73.12 | 2.29e-14 * |
| Gaming | 174.53 | < 2.2e-16 * |
| Social media use | 117.3 | < 2.2e-16 * |
| *Eating disorders* | | |
| TV watching | 8.73 | 0.12 |
| Gaming | 3.86 | 0.57 |
| Social media use | 29.54 | 1.81e-05 * |
| *Hyperkinetic disorders* | | |
| TV watching | 43.47 | 2.97e-08 * |
| Gaming | 148.83 | < 2.2e-16 * |
| Social media use | 84.34 | < 2.2e-16 * |
| *Pervasive developmental disorders* | | |
| TV watching | 27.76 | 4.06e-05 * |
| Gaming | 179.96 | < 2.2e-16 * |
| Social media use | 221.81 | < 2.2e-16 * |

*Original p-values are reported, with asterisks indicating statistically significant results after comparison to the family-wise error rate (α = 0.0033), adjusted using Bonferroni correction (n_tests_ = 15). Example of models compared: depressive_disorder_diagnosis ~ age + sex + socioeconomic_status vs. depressive_disorder_diagnosis ~ age + sex + socioeconomic_status + TV_watching_score.*

### Table S13. Results of likelihood ratio tests comparing full models (including screen use score) to baseline models (including only age and socioeconomic status) across specific diagnostic categories, stratified by sex assigned at birth (*n* = 21,598).

| **Phenotype** | **Females** | | **Males** | |
| --- | --- | --- | --- | --- |
|  | **ΔDeviance** | ***p­*-value** | **ΔDeviance** | ***p­*-value** |
| *Depressive disorders* | | |  |  |
| TV watching | 58.77 | 2.18e-11 * | 14.08 | 0.015 |
| Gaming | 76.95 | 3.65e-15 * | 34.01 | 2.38e-06 * |
| Social media use | 61.30 | 6.53e-12 * | 31.71 | 6.79e-06 * |
| *Anxiety/stress-related disorders* | | |  |  |
| TV watching | 68.30 | 2.32e-13 * | 19.09 | 0.0019 |
| Gaming | 132.23 | < 2.2e-16 * | 52.54 | 4.19e-10 * |
| Social media use | 80.50 | 6.58e-16 * | 39.26 | 2.11e-07 * |
| *Eating disorders* | | |  |  |
| TV watching | 9.07 | 0.11 | *NA* | *NA* |
| Gaming | 4.34 | 0.50 | *NA* | *NA* |
| Social media use | 27.17 | 5.29e-05 * | *NA* | *NA* |
| *Hyperkinetic disorders* | | |  |  |
| TV watching | 22.39 | 0.00044 * | 22.65 | 0.00039 * |
| Gaming | 95.59 | < 2.2e-16 * | 77.11 | 3.37e-15 * |
| Social media use | 42.24 | 5.26e-08 * | 44.07 | 2.24e-08 * |
| *Pervasive developmental disorders* | | |  |  |
| TV watching | 17.16 | 0.0042 | 12.24 | 0.032 |
| Gaming | 128.14 | < 2.2e-16 * | 78.45 | 1.77e-15 * |
| Social media use | 99.79 | < 2.2e-16 * | 127.20 | < 2.2e-16 * |

*Original p-values are reported, with asterisks indicating statistically significant results after comparison to the family-wise error rate (α = 0.0017), adjusted using Bonferroni correction (n_tests_ = 30). Analyses were conducted for diagnostic categories with a prevalence greater than 1% within each sex-specific subsample. “NA” indicates that the prevalence was below 1% in the respective subsample. Example of models compared: depressive_disorder_diagnosis_females ~ age + socioeconomic_status vs. depressive_disorder_diagnosis_females ~ age + socioeconomic_status + TV_watching_score.*

### Table S14. Results of the F-tests comparing full models (including screen use score) to baseline models (including only age, sex assigned at birth, and socioeconomic status), performed across specific scores representing self-reported symptoms of mental health problems in the subsample without any psychiatric diagnoses (*n* = 18,109).

| **Phenotype** | **F (df)** | ***p­*-value** |
| --- | --- | --- |
| *SMFQ score* | | |
| TV watching | 41.92 (5; 17,907) | < 2.2e-16 * |
| Gaming | 32.50 (5; 17,868) | < 2.2e-16 * |
| Social media use | 49.98 (5; 17,915) | < 2.2e-16 * |
| *SCARED score* | | |
| TV watching | 23.43 (5; 17,783) | < 2.2e-16 * |
| Gaming | 26.32 (5; 17,745) | < 2.2e-16 * |
| Social media use | 16.94 (5; 17,790) | < 2.2e-16 * |
| *EDE-Q score* | | |
| TV watching | 36.41 (5; 17,872) | < 2.2e-16 * |
| Gaming | 24.16 (5; 17,832) | < 2.2e-16 * |
| Social media use | 36.83 (5; 17,879) | < 2.2e-16 * |
| *RS-DBD score* | | |
| TV watching | 11.13 (5; 14,759) | 1.00e-10 * |
| Gaming | 25.84 (5; 14,724) | < 2.2e-16 * |
| Social media use | 30.02 (5; 14,764) | < 2.2e-16 * |
| *SCQ score* | | |
| TV watching | 5.86 (5; 13,685) | 2.07e-05 * |
| Gaming | 6.32 (5; 13,660) | 7.24e-06 * |
| Social media use | 2.70 (5; 13,692) | 0.019 |

*Original p-values are reported, with asterisks indicating statistically significant results after comparison to the family-wise error rate (α = 0.0033), adjusted using Bonferroni correction (n_tests_ = 15). Example of models compared: smfq_score ~ age + sex + socioeconomic_status vs. smfq_score ~ age + sex + socioeconomic_status + TV_watching_score.*

### Table S15. Results of the F-tests comparing full models (including screen use score) to baseline models (including only age and socioeconomic status) across specific diagnostic blocks/categories in the subsample without any psychiatric diagnoses (*n* = 18,109), stratified by sex assigned at birth.

| **Phenotype** | **F (df)** | ***p­*-value** | **F (df)** | ***p­*-value** |
| --- | --- | --- | --- | --- |
|  | **Females** | | **Males** | |
| *SFMQ score* | | | | |
| TV watching | 31.29 (5, 9419) | < 2.2e-16 * | 10.04 (5, 8481) | 1.34e-06 * |
| Gaming | 25.43 (5, 9364) | < 2.2e-16 * | 11.81 (5, 8497) | 2.07e-11 * |
| Social media use | 36.87 (5, 9414) | < 2.2e-16 * | 13.37 (5, 8494) | 5.24e-13 * |
| *SCARED score* | | | | |
| TV watching | 21.06 (5, 9390) | < 2.2e-16 * | 2.73 (5, 8386) | 0.018 |
| Gaming | 21.81 (5, 9334) | < 2.2e-16 * | 8.44 (5, 8404) | 5.62e-08 * |
| Social media use | 15.47 (5, 9384) | 3.48e-15 * | 3.82 (5, 8399) | 0.0018 |
| *EDE-Q score* | | | | |
| TV watching | 26.31 (5, 9402) | < 2.2e-16 * | 9.14 (5, 8436) | 1.11e-08 * |
| Gaming | 14.75 (5, 9345) | 1.95e-14 * | 16.43 (5, 8480) | 3.59e-16 * |
| Social media use | 25.99 (5, 9396) | < 2.2e-16 * | 11.04 (5, 8476) | 1.30e-10 * |
| *RS-DBD score* | | | | |
| TV watching | 6.98 (5, 7556) | 1.64e-06 * | 5.87 (5, 7196) | 2.04e-05 * |
| Gaming | 20.12 (5, 7513) | < 2.2e-16 * | 10.87 (5, 7204) | 1.95e-10 * |
| Social media use | 26.73 (5, 7551) | < 2.2e-16 * | 9.52 (5, 7206) | 4.64e-09 * |
| *SCQ score* | | | | |
| TV watching | 3.73 (5, 7018) | 0.0023 | 2.58 (5, 6660) | 0.025 |
| Gaming | 1.47 (5, 6982) | 0.20 | 5.57 (5, 6666) | 4.01e-05 * |
| Social media use | 2.89 (5, 7013) | 0.013 | 1.60 (5, 6672) | 0.16 |

*Original p-values are reported, with asterisks indicating statistically significant results after comparison to the family-wise error rate (α = 0.0017), adjusted using Bonferroni correction (n_tests_ = 30). Analyses were conducted for diagnostic categories with a prevalence greater than 1% within each sex-specific subsample. “NA” indicates that the prevalence was below 1% in the respective subsample. Example of models compared: smfq_score_females ~ age + socioeconomic_status vs. smfq_score_females ~ age + socioeconomic_status + TV_watching_score*

### Table S16. Results of the F-tests comparing full models (including screen use score) to baseline models (including only age, sex assigned at birth, and socioeconomic status), performed across specific scores representing self-reported symptoms of mental health problems in the subsample of participants with at least one psychiatric diagnosis registered (*n* = 3489).

| **Phenotype** | **F (df)** | ***p­*-value** |
| --- | --- | --- |
| *SMFQ score* | | |
| TV watching | 7.81 (5; 3436) | 2.5e-07 * |
| Gaming | 6.41 (5; 3437) | 6.13e-06 * |
| Social media use | 7.17 (5; 3446) | 1.10e-06 * |
| *SCARED score* | | |
| TV watching | 5.87 (5; 3414) | 2.07e-05 * |
| Gaming | 5.51 (5; 3414) | 4.63e-05 * |
| Social media use | 2.72 (5; 3423) | 0.018 |
| *EDE-Q score* | | |
| TV watching | 8.56 (5; 3423) | 4.60e-08 * |
| Gaming | 3.65 (5; 3424) | 0.0027 * |
| Social media use | 7.13 (5; 3433) | 1.20e-06 * |
| *RS-DBD score* | | |
| TV watching | 8.29 (5; 2712) | 8.69e-05 * |
| Gaming | 7.27 (5; 2714) | 8.89e-07 * |
| Social media use | 6.22 (5; 2722) | 9.72e-06 * |
| *SCQ score* | | |
| TV watching | 3.07 (5; 2460) | 0.0091 |
| Gaming | 5.14 (5; 2462) | 0.00011 * |
| Social media use | 6.93 (5; 2468) | 1.96e-06 * |

*Original p-values are reported, with asterisks indicating statistically significant results after comparison to the family-wise error rate (α = 0.0033), adjusted using Bonferroni correction (n_tests_ = 15). Example of models compared: smfq_score_subsample_with_diagnoses ~ age + sex + socioeconomic_status vs. smfq_score_subsample_with_diagnoses ~ age + sex + socioeconomic_status + TV_watching_score.*

### Table S17. Results of likelihood ratio tests comparing full models (including polygenic risk score for a given psychiatric disorder) to baseline models (including only age, genetic sex, and socioeconomic status), performed across diagnostic categories with a prevalence greater than 1% in the study sample.

| **Screen-based behavior** | **PRS** | **ΔDeviance** | ***p­*-value** |
| --- | --- | --- | --- |
| TV watching | MD | 26.48 | 7.21e-05 * |
|  | ANX | 9.33 | 0.096 |
|  | AN | 14.91 | 0.011 |
|  | ADHD | 34.25 | 2.13e-06 * |
|  | ASD | 6.15 | 0.29 |
| Gaming | MD | 20.58 | 0.00097 * |
|  | ANX | 3.66 | 0.60 |
|  | AN | 32.25 | 5.30e-05 * |
|  | ADHD | 51.76 | 6.04e-10 * |
|  | ASD | 29.58 | 1.78e-05 * |
| Social media use | MD | 43.76 | 2.60e-08 * |
|  | ANX | 3.29 | 0.65 |
|  | AN | 36.01 | 9.44e-07 * |
|  | ADHD | 91.16 | < 2.2e-16 * |
|  | ASD | 41.36 | 7.93e-08 * |

*Original p-values are reported, with asterisks indicating statistically significant results after comparison to the family-wise error rate (α = 0.0033), adjusted using Bonferroni correction (n_tests_ = 15). Example of models compared: gaming_score ~ age + sex + socioeconomic_status vs. gaming_score ~ age + sex + socioeconomic_status + PRS_ADHD_preresidualized.*

*MD: major depression; ANX: anxiety disorder; AN: anorexia nervosa; ADHD: attention-deficit hyperactivity disorder; ASD: autism spectrum disorder; PRS: polygenic risk score*

### Table S18. Results of likelihood ratio tests comparing full models (including polygenic risk score for a given psychiatric disorder) to baseline models (including only age, genetic sex, and socioeconomic status), performed across diagnostic categories with a prevalence lower than 1% in the study sample.

| **Screen-based behavior** | **PRS** | **ΔDeviance** | ***p­*-value** |
| --- | --- | --- | --- |
| TV watching | SCZ | 6.17 | 0.29 |
|  | BP | 7.64 | 0.18 |
|  | AUD | 4.24 | 0.52 |
| Gaming | SCZ | 41.02 | 9.30e-08 * |
|  | BP | 18.39 | 0.0025 * |
|  | AUD | 2.68 | 0.75 |
| Social media use | SCZ | 25.69 | 0.00010 * |
|  | BP | 12.34 | 0.030 |
|  | AUD | 18.37 | 0.0025 * |

*Original p-values are reported, with asterisks indicating statistically significant results after comparison to the family-wise error rate (α = 0.0056), adjusted using Bonferroni correction (n_tests_ = 9). Example of models compared: gaming_score ~ age + sex + socioeconomic_status vs. gaming_score ~ age + sex + socioeconomic_status + PRS_SCZ_preresidualized.*

*SCZ: schizophrenia; BP: bipolar disorder; AUD: alcohol use disorder; PRS: polygenic risk score*

### Table S19. Results of likelihood ratio tests comparing full models (including polygenic risk score for a given psychiatric disorder) to baseline models (including only age, genetic sex, and socioeconomic status), performed across diagnostic categories with a prevalence greater than 1% in the study sample, within a subsample of participants without any psychiatric diagnosis registered (*n* = 14,625).

| **Screen-based behavior** | **PRS** | **ΔDeviance** | ***p­*-value** |
| --- | --- | --- | --- |
| TV watching | MD | 14.10 | 0.015 |
|  | ANX | 6.81 | 0.24 |
|  | AN | 18.90 | 0.0020 * |
|  | ADHD | 18.35 | 0.0025 * |
|  | ASD | 6.62 | 0.25 |
| Gaming | MD | 11.05 | 0.05 |
|  | ANX | 4.01 | 0.55 |
|  | AN | 26.28 | 7.87e-05 * |
|  | ADHD | 34.80 | 1.65e-06 * |
|  | ASD | 28.84 | 2.49e-05 * |
| Social media use | MD | 27.53 | 4.50e-05 * |
|  | ANX | 2.62 | 0.76 |
|  | AN | 31.13 | 8.83e-06 * |
|  | ADHD | 64.00 | 1.80e-12 * |
|  | ASD | 30.16 | 1.37e-05 * |

*Original p-values are reported, with asterisks indicating statistically significant results after comparison to the family-wise error rate (α = 0.0033), adjusted using Bonferroni correction (n_tests_ = 15). Example of models compared: gaming_score_healthy_subsample ~ age + sex + socioeconomic_status vs. gaming_score_healthy_subsample ~ age + sex + socioeconomic_status + PRS_ADHD_preresidualized.*

*MD: major depression; ANX: anxiety disorder; AN: anorexia nervosa; ADHD: attention-deficit hyperactivity disorder; ASD: autism spectrum disorder; PRS: polygenic risk score*

### Table S20. Full model output for Gsens sensitivity analyses (moderate-to-high TV watching, female subsample, *n* = 6598).

|  | **est** | **se** | **z** | **p-value** | **ci.lower** | **ci.upper** | **scenario** | **phenotype** |
| --- | --- | --- | --- | --- | --- | --- | --- | --- |
| Adjusted Bxy | 0.075 | 0.012 | 6.132 | 8.68e-10 * | 0.051 | 0.1 | base | ADHD |
| Genetic confounding | 0.014 | 0.002 | 6.511 | 7.47e-11 * | 0.009 | 0.018 |  |  |
| Total effect | 0.089 | 0.012 | 7.256 | 3.98e-13 * | 0.065 | 0.113 |  |  |
| Adjusted Bxy | 0.021 | 0.016 | 1.348 | 0.1775 | -0.01 | 0.052 | snp |  |
| Genetic confounding | 0.068 | 0.011 | 6.161 | 7.24e-10 * | 0.046 | 0.089 |  |  |
| Total effect | 0.089 | 0.012 | 7.224 | 5.04e-13 * | 0.065 | 0.113 |  |  |
| Adjusted Bxy | 0.0 | 0.0 | NA | NA | 0.0 | 0.0 | twin |  |
| Genetic confounding | 0.094 | 0.012 | 7.588 | 3.25e-14 * | 0.069 | 0.118 |  |  |
| Total effect | 0.094 | 0.012 | 7.588 | 3.25e-14 * | 0.069 | 0.118 |  |  |
| Adjusted Bxy | 0.122 | 0.012 | 10.169 | 2.73e-24 * | 0.099 | 0.146 | base | MD |
| Genetic confounding | 0.008 | 0.002 | 3.352 | 0.00080 * | 0.003 | 0.012 |  |  |
| Total effect | 0.13 | 0.012 | 10.649 | 1.77e-26 * | 0.106 | 0.154 |  |  |
| Adjusted Bxy | 0.108 | 0.013 | 8.416 | 3.89e-17 * | 0.083 | 0.133 | snp |  |
| Genetic confounding | 0.022 | 0.007 | 3.332 | 0.00086 * | 0.009 | 0.035 |  |  |
| Total effect | 0.13 | 0.012 | 10.644 | 1.86e-26 * | 0.106 | 0.154 |  |  |
| Adjusted Bxy | 0.057 | 0.024 | 2.416 | 0.016 | 0.011 | 0.103 | twin |  |
| Genetic confounding | 0.073 | 0.022 | 3.263 | 0.0011 * | 0.029 | 0.117 |  |  |
| Total effect | 0.13 | 0.012 | 10.592 | 3.26e-26 * | 0.106 | 0.154 |  |  |

*Adjusted_Bxy: estimate of the relationship between moderate-to-high TV watching and diagnosis in the female subsample after accounting for the polygenic risk score (PRS) under three scenarios (observed PRS [base], PRS that explains SNP-heritability [snp], and PRS that explains twin heritability [twin]); Genetic confounding: estimate of genetic confounding; Total effect: total observed association.*

*ADHD: attention-deficit hyperactivity disorder; MD: major depression*

*Original p-values are reported, with asterisks indicating statistically significant results after comparison to the family-wise error rate (α = 0.0083), adjusted using Bonferroni correction (n_tests_ = 6).*

### Table S21. Full model output for Gsens sensitivity analyses (moderate-to-high TV watching, male subsample, *n* = 4356).

|  | **est** | **se** | **z** | **p-value** | **ci.lower** | **ci.upper** | **scenario** | **phenotype** |
| --- | --- | --- | --- | --- | --- | --- | --- | --- |
| Adjusted Bxy | 0.092 | 0.015 | 6.183 | 6.29e-10 * | 0.063 | 0.121 | base | ADHD |
| Genetic confounding | 0.008 | 0.003 | 2.89 | 0.0039 * | 0.003 | 0.013 |  |  |
| Total effect | 0.1 | 0.015 | 6.632 | 3.31e-11 * | 0.07 | 0.13 |  |  |
| Adjusted Bxy | 0.069 | 0.017 | 4.024 | 5.73e-05 * | 0.035 | 0.102 | snp |  |
| Genetic confounding | 0.031 | 0.011 | 2.859 | 0.0043 * | 0.01 | 0.053 |  |  |
| Total effect | 0.1 | 0.015 | 6.626 | 3.45e-11 * | 0.07 | 0.13 |  |  |
| Adjusted Bxy | 0.0 | 0.0 | NA | NA | 0.0 | 0.0 | twin |  |
| Genetic confounding | 0.101 | 0.015 | 6.674 | 2.49e-11 * | 0.072 | 0.131 |  |  |
| Total effect | 0.101 | 0.015 | 6.674 | 2.49e-11 * | 0.072 | 0.131 |  |  |
| Adjusted Bxy | 0.104 | 0.015 | 6.86 | 6.91e-12 * | 0.074 | 0.134 | base | MD |
| Genetic confounding | 0.016 | 0.003 | 6.17 | 6.82e-10 * | 0.011 | 0.021 |  |  |
| Total effect | 0.12 | 0.015 | 7.975 | 1.53e-15 * | 0.091 | 0.149 |  |  |
| Adjusted Bxy | 0.062 | 0.017 | 3.564 | 0.00037 * | 0.028 | 0.097 | snp |  |
| Genetic confounding | 0.058 | 0.01 | 5.882 | 4.05e-09 * | 0.038 | 0.077 |  |  |
| Total effect | 0.12 | 0.015 | 7.95 | 1.87e-15 * | 0.09 | 0.15 |  |  |
| Adjusted Bxy | 0.0 | 0.0 | NA | NA | 0.0 | 0.0 | twin |  |
| Genetic confounding | 0.132 | 0.014 | 9.187 | 4.05e-20 * | 0.104 | 0.16 |  |  |
| Total effect | 0.132 | 0.014 | 9.187 | 4.05e-20 * | 0.104 | 0.16 |  |  |

*Adjusted_Bxy: estimate of the relationship between moderate-to-high TV watching and diagnosis in the male subsample after accounting for the polygenic risk score (PRS) under three scenarios (observed PRS [base], PRS that explains SNP-heritability [snp], and PRS that explains twin heritability [twin]); Genetic confounding: estimate of genetic confounding; Total effect: total observed association.*

*ADHD: attention-deficit hyperactivity disorder; MD: major depression*

*Original p-values are reported, with asterisks indicating statistically significant results after comparison to the family-wise error rate (α = 0.0083), adjusted using Bonferroni correction (n_tests_ = 6).*

### Table S22. Full model output for Gsens sensitivity analyses (moderate-to-high gaming, female subsample, *n* = 2204).

|  | **est** | **se** | **z** | **p-value** | **ci.lower** | **ci.upper** | **scenario** | **phenotype** |
| --- | --- | --- | --- | --- | --- | --- | --- | --- |
| Adjusted Bxy | 0.188 | 0.021 | 8.999 | 2.27e-19 * | 0.147 | 0.229 | base | ASD |
| Genetic confounding | 0.002 | 0.001 | 1.526 | 0.13 | 0.0 | 0.004 |  |  |
| Total effect | 0.19 | 0.021 | 9.083 | 1.05e-19 * | 0.149 | 0.231 |  |  |
| Adjusted Bxy | 0.137 | 0.042 | 3.242 | 0.0012 * | -0.01 | 0.052 | snp |  |
| Genetic confounding | 0.053 | 0.037 | 1.439 | 0.15027 | 0.046 | 0.089 |  |  |
| Total effect | 0.19 | 0.021 | 9.057 | 1.35e-19 * | 0.065 | 0.113 |  |  |
| Adjusted Bxy | 0.0 | 0.0 | NA | NA | 0.0 | 0.0 | twin |  |
| Genetic confounding | 0.19 | 0.022 | 8.778 | 1.66e-18 * | 0.148 | 0.233 |  |  |
| Total effect | 0.19 | 0.022 | 8.778 | 1.66e-18 * | 0.148 | 0.233 |  |  |

*Adjusted_Bxy: estimate of the relationship between moderate-to-high gaming and diagnosis in the female subsample after accounting for the polygenic risk score (PRS) under three scenarios (observed PRS [base], PRS that explains SNP-heritability [snp], and PRS that explains twin heritability [twin]); Genetic confounding: estimate of genetic confounding; Total effect: total observed association.*

*ASD: autism spectrum disorder*

*Original p-values are reported, with asterisks indicating statistically significant results after comparison to the family-wise error rate (α = 0.017), adjusted using Bonferroni correction (n_tests_ = 3).*

### Table S23. Full model output for Gsens sensitivity analyses (moderate-to-high gaming, male subsample, *n* = 6247).

|  | **est** | **se** | **z** | **p-value** | **ci.lower** | **ci.upper** | **scenario** | **phenotype** |
| --- | --- | --- | --- | --- | --- | --- | --- | --- |
| Adjusted Bxy | 0.182 | 0.012 | 14.808 | 1.31e-49 * | 0.158 | 0.207 | base | ADHD |
| Genetic confounding | 0.008 | 0.002 | 3.979 | 6.91e-05 * | 0.004 | 0.011 |  |  |
| Total effect | 0.19 | 0.012 | 15.294 | 8.42e-53 * | 0.166 | 0.214 |  |  |
| Adjusted Bxy | 0.153 | 0.015 | 10.477 | 1.10e-25 * | 0.124 | 0.182 | snp |  |
| Genetic confounding | 0.037 | 0.009 | 3.908 | 9.29e-05 * | 0.018 | 0.056 |  |  |
| Total effect | 0.19 | 0.012 | 15.273 | 1.16e-52 * | 0.166 | 0.214 |  |  |
| Adjusted Bxy | 0.0 | 0.0 | NA | NA | 0.0 | 0.0 | twin |  |
| Genetic confounding | 0.19 | 0.013 | 14.822 | 1.05e-49 * | 0.165 | 0.215 |  |  |
| Total effect | 0.19 | 0.013 | 14.822 | 1.05e-49 * | 0.165 | 0.215 |  |  |
| Adjusted Bxy | 0.154 | 0.012 | 12.456 | 1.30e-35 * | 0.13 | 0.178 | base | MD |
| Genetic confounding | 0.006 | 0.002 | 3.063 | 0.0022 * | 0.002 | 0.01 |  |  |
| Total effect | 0.16 | 0.012 | 12.81 | 1.45e-37 * | 0.136 | 0.184 |  |  |
| Adjusted Bxy | 0.136 | 0.014 | 9.904 | 4.00e-23 * | 0.109 | 0.163 | snp |  |
| Genetic confounding | 0.024 | 0.008 | 3.038 | 0.0024 * | 0.008 | 0.039 |  |  |
| Total effect | 0.16 | 0.012 | 12.803 | 1.59e-37 * | 0.136 | 0.184 |  |  |
| Adjusted Bxy | 0.081 | 0.028 | 2.915 | 0.0036 * | 0.027 | 0.135 | twin |  |
| Genetic confounding | 0.079 | 0.027 | 2.962 | 0.0031 * | 0.027 | 0.131 |  |  |
| Total effect | 0.16 | 0.013 | 12.729 | 4.10e-37 * | 0.135 | 0.185 |  |  |
| Adjusted Bxy | 0.226 | 0.012 | 18.405 | 1.19e-75 * | 0.202 | 0.25 |  | ASD |
| Genetic confounding | 0.004 | 0.001 | 3.237 | 0.0012 * | 0.002 | 0.007 |  |  |
| Total effect | 0.23 | 0.012 | 18.678 | 7.52e-78 * | 0.206 | 0.254 |  |  |
| Adjusted Bxy | 0.196 | 0.016 | 12.504 | 7.07e-36 * | 0.165 | 0.227 |  |  |
| Genetic confounding | 0.034 | 0.011 | 3.169 | 0.0015 * | 0.013 | 0.055 |  |  |
| Total effect | 0.23 | 0.012 | 18.655 | 1.15e-77 * | 0.206 | 0.254 |  |  |
| Adjusted Bxy | 0.0 | 0.0 | NA | NA | 0.0 | 0.0 |  |  |
| Genetic confounding | 0.23 | 0.013 | 17.743 | 1.94e-70 * | 0.205 | 0.256 |  |  |
| Total effect | 0.23 | 0.013 | 17.743 | 1.94e-70 * | 0.205 | 0.256 |  |  |

*Adjusted_Bxy: estimate of the relationship between moderate-to-high gaming and diagnosis in the male subsample after accounting for the polygenic risk score (PRS) under three scenarios (observed PRS [base], PRS that explains SNP-heritability [snp], and PRS that explains twin heritability [twin]); Genetic confounding: estimate of genetic confounding; Total effect: total observed association.*

*ADHD: attention-deficit hyperactivity disorder; MD: major depression; ASD: autism spectrum disorder*

*Original p-values are reported, with asterisks indicating statistically significant results after comparison to the family-wise error rate (α = 0.0056), adjusted using Bonferroni correction (n_tests_ = 9).*

### Table S24. Full model output for Gsens sensitivity analyses (moderate-to-high social media use, female subsample, *n* = 7270).

|  | **est** | **se** | **z** | **p-value** | **ci.lower** | **ci.upper** | **scenario** | **phenotype** |
| --- | --- | --- | --- | --- | --- | --- | --- | --- |
| Adjusted Bxy | 0.105 | 0.012 | 8.947 | 3.66e-19 * | 0.082 | 0.128 | base | ADHD |
| Genetic confounding | 0.015 | 0.002 | 7.844 | 4.38e-15 * | 0.011 | 0.019 |  |  |
| Total effect | 0.12 | 0.012 | 10.303 | 6.82e-25 * | 0.097 | 0.143 |  |  |
| Adjusted Bxy | 0.044 | 0.015 | 2.923 | 0.0035 * | 0.015 | 0.074 | snp |  |
| Genetic confounding | 0.076 | 0.01 | 7.294 | 3.00e-13 * | 0.055 | 0.096 |  |  |
| Total effect | 0.12 | 0.012 | 10.246 | 1.23e-24 * | 0.097 | 0.143 |  |  |
| Adjusted Bxy | 0.0 | 0.0 | NA | NA | 0.0 | 0.0 | twin |  |
| Genetic confounding | 0.125 | 0.012 | 10.637 | 2.00e-26 * | 0.102 | 0.148 |  |  |
| Total effect | 0.125 | 0.012 | 10.637 | 2.00e-26 * | 0.102 | 0.148 |  |  |
| Adjusted Bxy | 0.094 | 0.012 | 8.125 | 4.46e-16 * | 0.072 | 0.117 | base | MD |
| Genetic confounding | 0.016 | 0.002 | 6.922 | 4.45e-12 * | 0.011 | 0.02 |  |  |
| Total effect | 0.11 | 0.012 | 9.433 | 3.97e-21 * | 0.087 | 0.133 |  |  |
| Adjusted Bxy | 0.065 | 0.013 | 5.148 | 2.63e-07 * | 0.041 | 0.09 | snp |  |
| Genetic confounding | 0.045 | 0.007 | 6.751 | 1.47e-11 * | 0.032 | 0.057 |  |  |
| Total effect | 0.11 | 0.012 | 9.417 | 4.65e-21 * | 0.087 | 0.133 |  |  |
| Adjusted Bxy | 0.0 | 0.0 | NA | NA | 0.0 | 0.0 | twin |  |
| Genetic confounding | 0.117 | 0.011 | 10.637 | 2.01e-26 * | 0.096 | 0.139 |  |  |
| Total effect | 0.117 | 0.011 | 10.637 | 2.01e-26 * | 0.096 | 0.139 |  |  |

*Adjusted_Bxy: estimate of the relationship between moderate-to-high social media use and diagnosis in the female subsample after accounting for the polygenic risk score (PRS) under three scenarios (observed PRS [base], PRS that explains SNP-heritability [snp], and PRS that explains twin heritability [twin]); Genetic confounding: estimate of genetic confounding; Total effect: total observed association.*

*ADHD: attention-deficit hyperactivity disorder; MD: major depression*

*Original p-values are reported, with asterisks indicating statistically significant results after comparison to the family-wise error rate (α = 0.0083), adjusted using Bonferroni correction (n_tests_ = 6).*

### Table S25. Full model output for Gsens sensitivity analyses (moderate-to-high social media use, male subsample, *n* = 5551).

|  | **est** | **se** | **z** | **p-value** | **ci.lower** | **ci.upper** | **scenario** | **phenotype** |
| --- | --- | --- | --- | --- | --- | --- | --- | --- |
| Adjusted Bxy | 0.12 | 0.013 | 9.126 | 7.13e-20 * | 0.095 | 0.146 | base | ADHD |
| Genetic confounding | 0.01 | 0.002 | 4.229 | 2.35e-05 * | 0.005 | 0.014 |  |  |
| Total effect | 0.13 | 0.013 | 9.767 | 1.56e-22 * | 0.104 | 0.156 |  |  |
| Adjusted Bxy | 0.088 | 0.016 | 5.679 | 1.35e-08 * | 0.058 | 0.119 | snp |  |
| Genetic confounding | 0.042 | 0.01 | 4.141 | 3.46e-05 * | 0.022 | 0.061 |  |  |
| Total effect | 0.13 | 0.013 | 9.751 | 1.84e-22 * | 0.104 | 0.156 |  |  |
| Adjusted Bxy | 0.0 | 0.0 | NA | NA | 0.0 | 0.0 | twin |  |
| Genetic confounding | 0.132 | 0.013 | 9.77 | 1.51e-22 * | 0.105 | 0.158 |  |  |
| Total effect | 0.132 | 0.013 | 9.77 | 1.51e-22 * | 0.105 | 0.158 |  |  |
| Adjusted Bxy | 0.089 | 0.013 | 6.775 | 1.24e-11 * | 0.063 | 0.115 | base | MD |
| Genetic confounding | 0.009 | 0.003 | 3.411 | 0.00065 * | 0.004 | 0.014 |  |  |
| Total effect | 0.098 | 0.013 | 7.336 | 2.21e-13 * | 0.072 | 0.124 |  |  |
| Adjusted Bxy | 0.073 | 0.014 | 5.175 | 2.28e-07 * | 0.045 | 0.101 | snp |  |
| Genetic confounding | 0.025 | 0.007 | 3.385 | 0.00071 * | 0.011 | 0.039 |  |  |
| Total effect | 0.098 | 0.013 | 7.332 | 2.28e-13 * | 0.072 | 0.124 |  |  |
| Adjusted Bxy | 0.015 | 0.026 | 0.571 | 0.57 | -0.037 | 0.067 | twin |  |
| Genetic confounding | 0.083 | 0.025 | 3.296 | 0.00098 * | 0.034 | 0.132 |  |  |
| Total effect | 0.098 | 0.013 | 7.286 | 3.20e-13 * | 0.072 | 0.124 |  |  |

*Adjusted_Bxy: estimate of the relationship between moderate-to-high social media use and diagnosis in the male subsample after accounting for the polygenic risk score (PRS) under three scenarios (observed PRS [base], PRS that explains SNP-heritability [snp], and PRS that explains twin heritability [twin]); Genetic confounding: estimate of genetic confounding; Total effect: total observed association.*

*ADHD: attention-deficit hyperactivity disorder; MD: major depression*

*Original p-values are reported, with asterisks indicating statistically significant results after comparison to the family-wise error rate (α = 0.0083), adjusted using Bonferroni correction (n_tests_ = 6).*

### Supplementary Figures

### Figure S1. Flow diagram for the study sample.

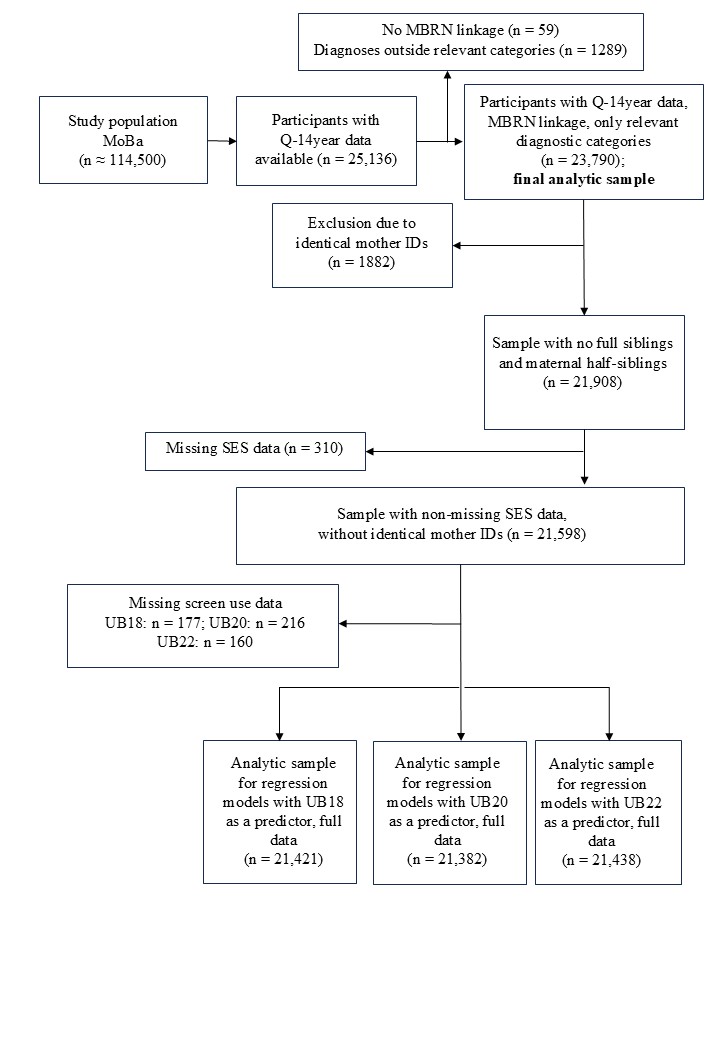

### Figure S2. Distribution of scores for the screen-based activities in the final analytic sample (*n* = 23,790).

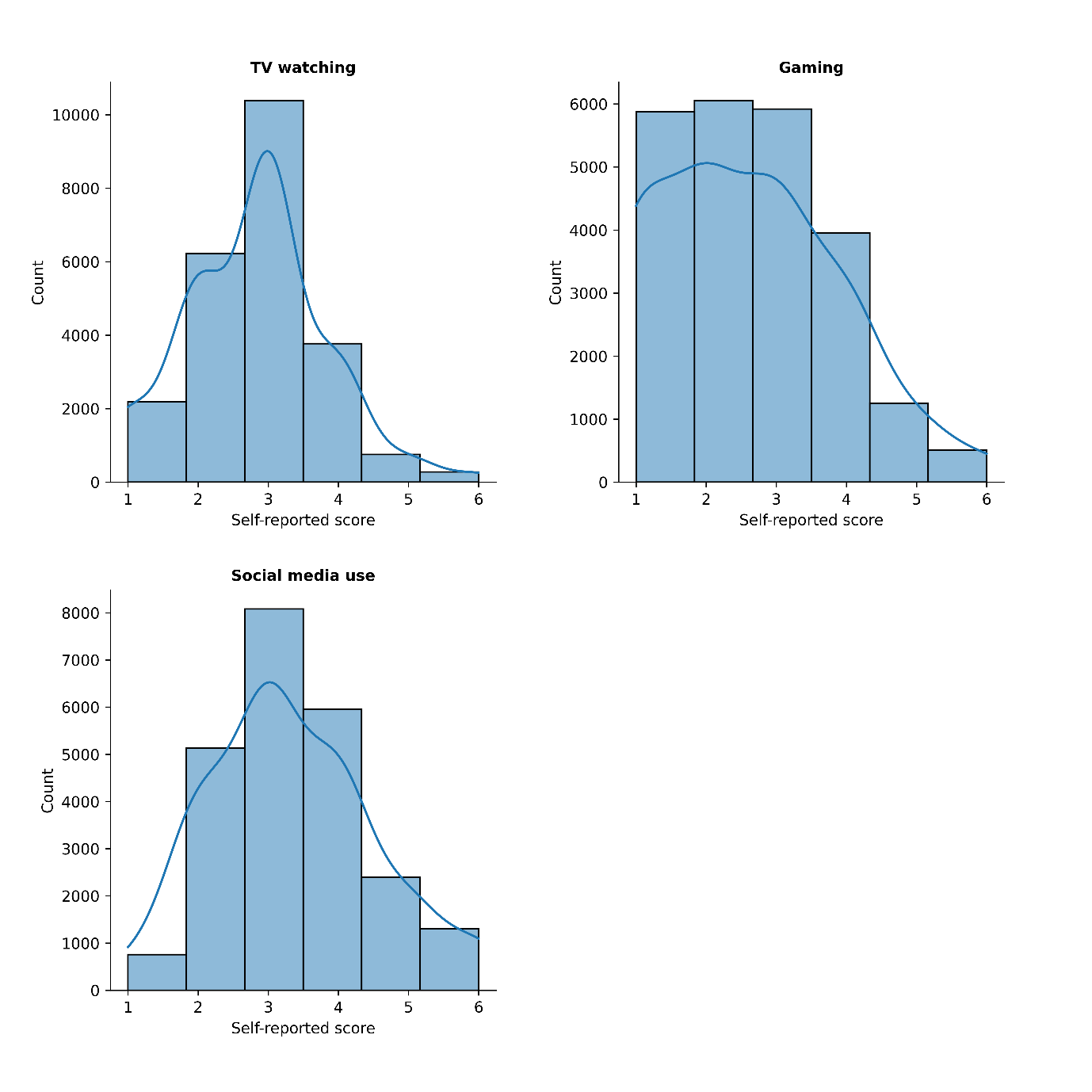

*Self-reported scores correspond to response options in the Q14-year questionnaire, and reflect time spent on a specific screen-based activity: 1: never/rarely; 2: less than 1 hour; 3: 1-2 hours; 4: 3-4 hours; 5: 5-6 hours; 6: 7 hours or more.*

### Figure S3. Distribution of scores for the screen-based activities in the final analytic sample (*n* = 23,790), stratified by sex assigned at birth.

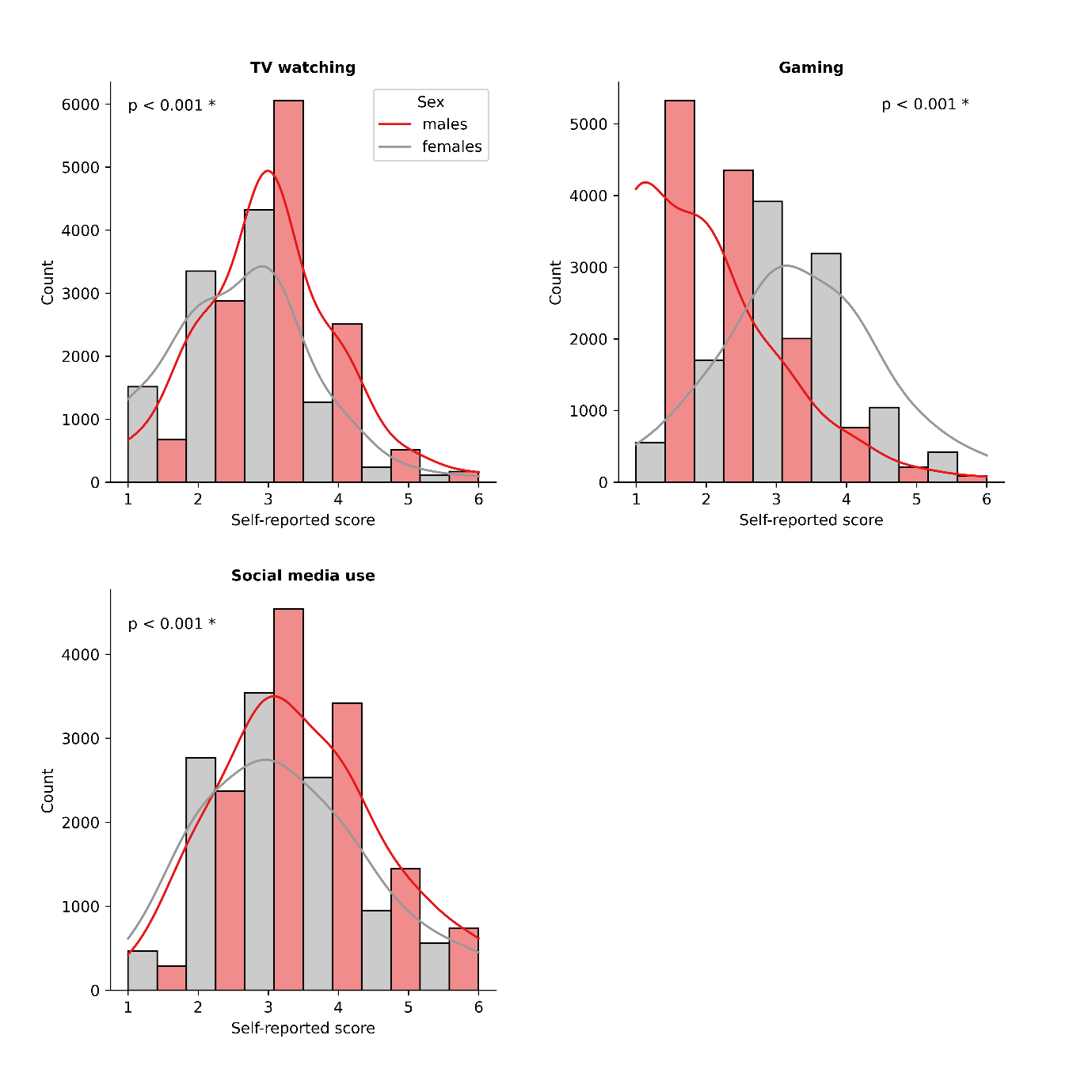

*Self-reported scores correspond to response options in the Q14-year questionnaire, and reflect time spent on a specific screen-based activity: 1: never/rarely; 2: less than 1 hour; 3: 1-2 hours; 4: 3-4 hours; 5: 5-6 hours; 6: 7 hours or more.*

*Comparison of score distributions in females and males was performed using Mann Witney U test, with asterisks indicating statistically significant results after comparison to the family-wise error rate (α = 0.017), adjusted using Bonferroni correction (n_tests_ = 3).*

### Figure S4. Odds ratios of having any psychiatric diagnosis among participants with different levels of screen-based activities, stratified by sex assigned at birth (*n_females_* = 11,742, *n_males_* = 9856).

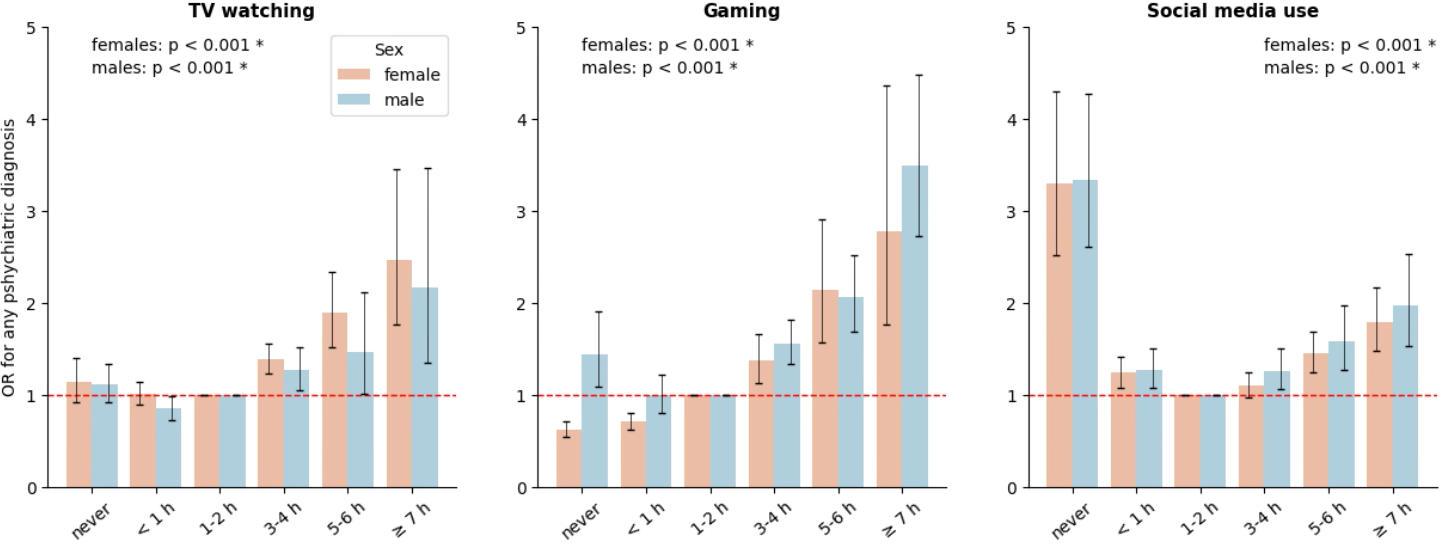

*Odds ratios of having any psychiatric diagnosis (within the selected diagnostic blocks/categories) among females and males with different levels of screen-based activities were estimated using logistic regression models with age and socioeconomic status as covariates. Level “1–2 hours per day” was used as a reference. P-values from the likelihood ratio tests comparing models with and without screen use variable are reported above each panel, with asterisks indicating statistically significant results after comparison to the family-wise error rate (α = 0.0083), adjusted using Bonferroni correction (n_tests_ = 6). The horizontal line indicates no association (OR = 1). Error bars represent 95% confidence intervals.*

### Figure S5. Odds ratios of having a psychiatric diagnosis among participants with different levels of screen-based activities, restricted to diagnoses received one year or later after Q14-year completion (*n* = 18,976).

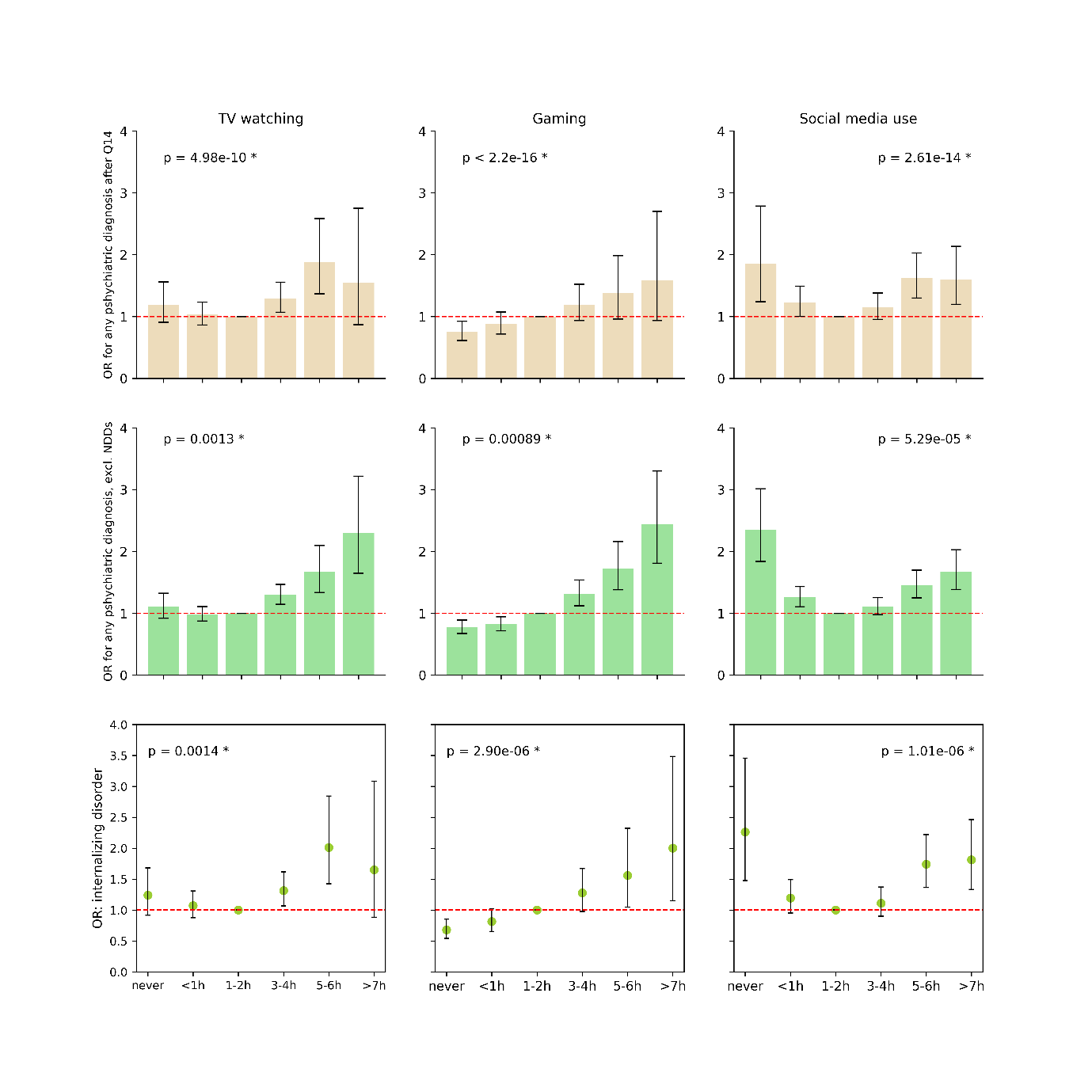

*The upper panel displays associations between screen-based activities and lifetime diagnoses of psychiatric disorders within relevant categories, excluding hyperkinetic and pervasive developmental disorders. The middle panel displays associations with the same diagnostic categories, restricted to diagnoses received one year or later after Q14-year completion. The lower panel displays associations between screen-based activities and internalizing disorder diagnoses received one year or later after Q14 completion. Internalizing disorders included F32-F34.1 (depressive disorders), and F40-F43, F93.0, F93.1, F93.2 (anxiety and stress-related disorders).*

*Odds ratios of having psychiatric diagnoses among participants with different levels of screen-based activities were estimated using logistic regression models with age, sex and socioeconomic status as covariates. Level “1–2 hours per day” was used as a reference.*

*P-values from the likelihood ratio tests comparing models with and without screen use level variable are reported above each panel, with asterisks indicating statistically significant results after comparison to the family-wise error rate (α = 0.017), adjusted using Bonferroni correction (n_tests_ = 3). The horizontal line indicates no association (OR = 1). Error bars represent 95% confidence intervals.*

### Figure S6. Associations between screen-based activities and lifetime psychiatric diagnoses across specific diagnostic categories, stratified by sex assigned at birth (*n* = 21,598).

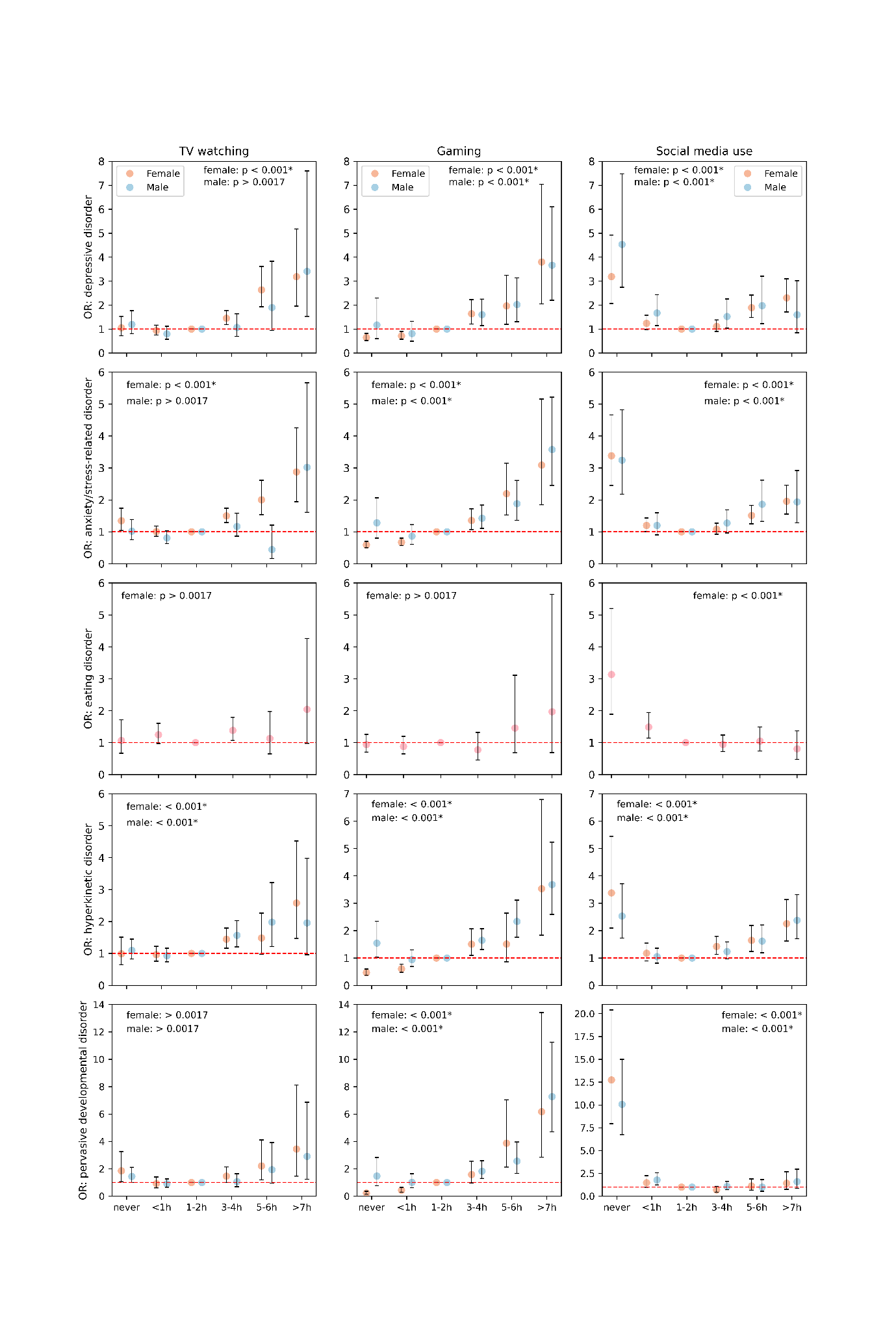

*Odds ratios of having a psychiatric diagnosis (within specific diagnostic category) among participants with different levels of screen-based activities were estimated using logistic regression models with age and socioeconomic status as covariates. The “1–2 hours per day” group was used as a reference. Analyses were conducted for diagnostic categories with a prevalence greater than 1% within each sex-specific subsample. P-values from the likelihood ratio tests comparing models with and without screen use variable are reported above each panel, with asterisks indicating statistically significant results after comparison to the family-wise error rate (α = 0.0017), adjusted using Bonferroni correction (n_tests_ = 30). The horizontal line indicates no association (OR = 1). Error bars represent 95% confidence intervals.*

### Figure S7. Associations between screen-based activities and standardized symptom scores among participants without psychiatric diagnoses (*n* = 18,109), stratified by sex assigned at birth.

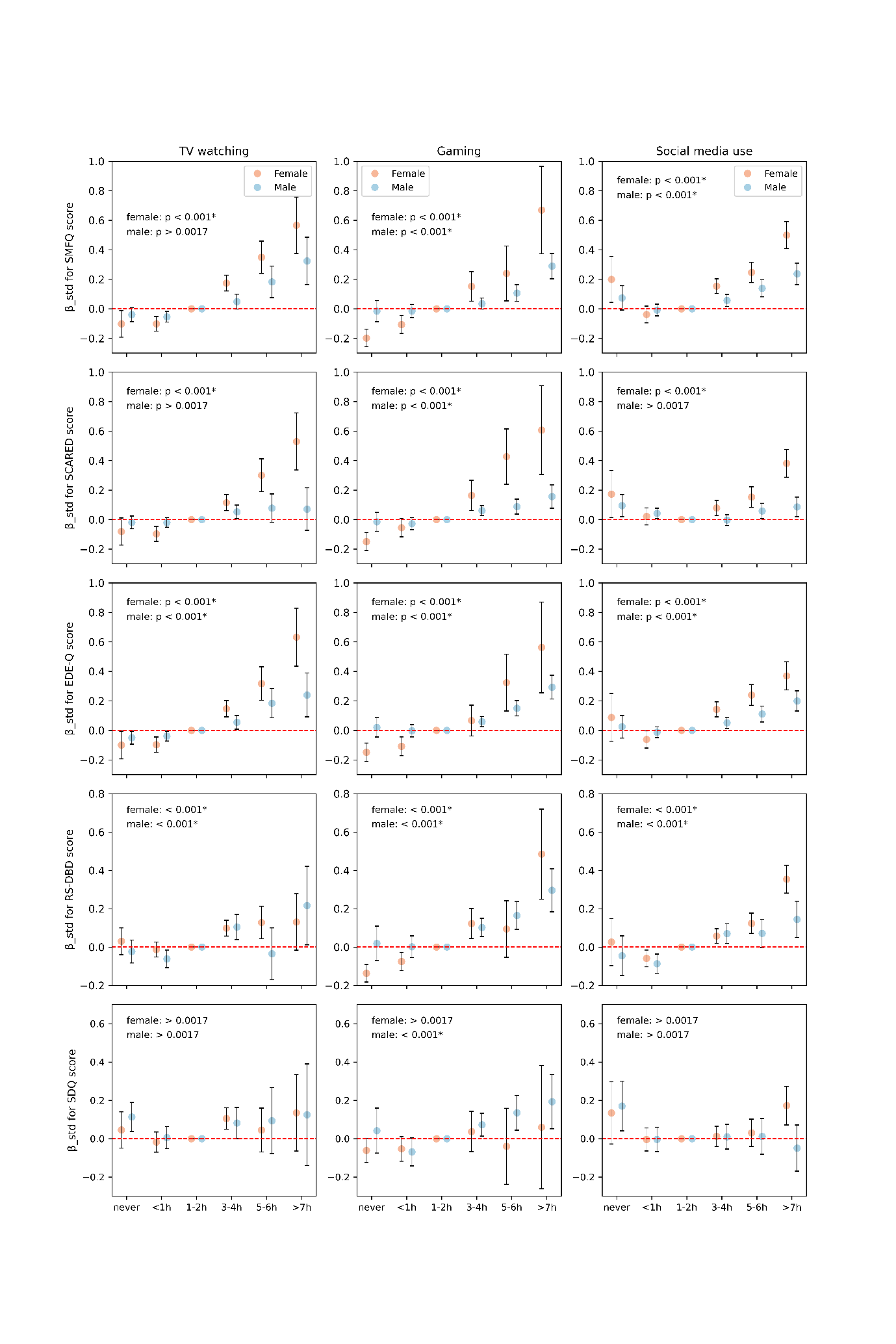

*Associations between levels of screen-based activities and standardized symptom scores were estimated using linear regression models with age and socioeconomic status as covariates. The “1–2 hours per day” group was used as a reference. P-values from F-tests comparing models with and without screen use variable are reported above each panel, with asterisks indicating statistically significant results after comparison to the family-wise error rate (α = 0.0017), adjusted using Bonferroni correction (n_tests_ = 30). The horizontal line indicates no difference from the reference group (β = 0). Error bars represent 95% confidence intervals.*

### Figure S8. Associations between levels of screen-based activities and standardized symptom scores among participants without psychiatric diagnoses (*n* = 18,109), and with at least one psychiatric diagnosis registered (*n* = 3489).

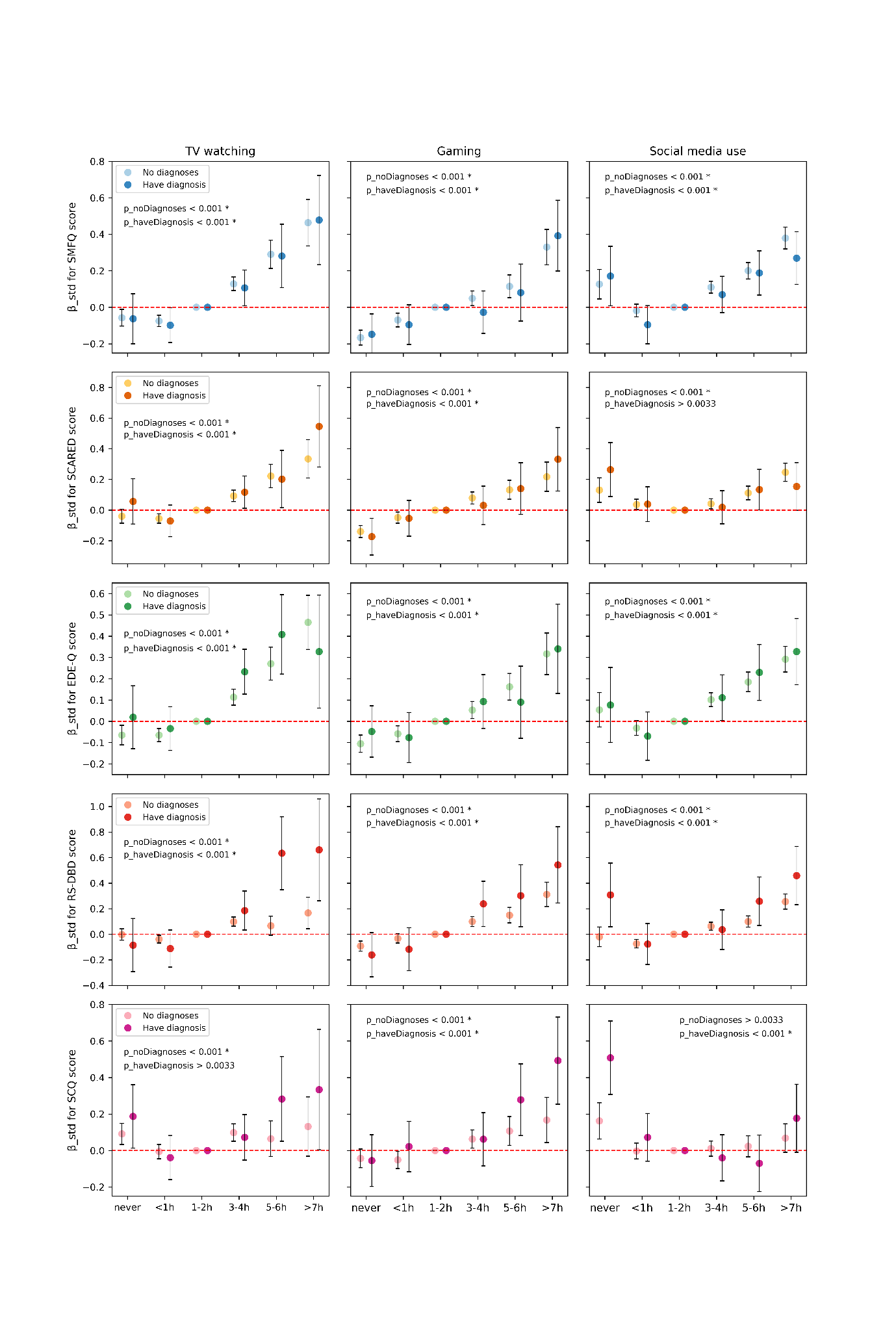

*Associations between levels of screen-based activities and standardized symptom scores were estimated using linear regression models with age, sex, and socioeconomic status as covariates. The “1–2 hours per day” group was used as a reference. P-values from F-tests comparing models with and without screen use variable are reported above each panel, with asterisks indicating statistically significant results after comparison to the family-wise error rate (α = 0.0017), adjusted using Bonferroni correction (n_tests_ = 30). The horizontal line indicates no difference from the reference group (β = 0). Error bars represent 95% confidence intervals.*

### Figure S9. Associations between levels of screen-based activities and polygenic risk scores for schizophrenia, bipolar disorder, and alcohol use disorder (*n* = 17,214).

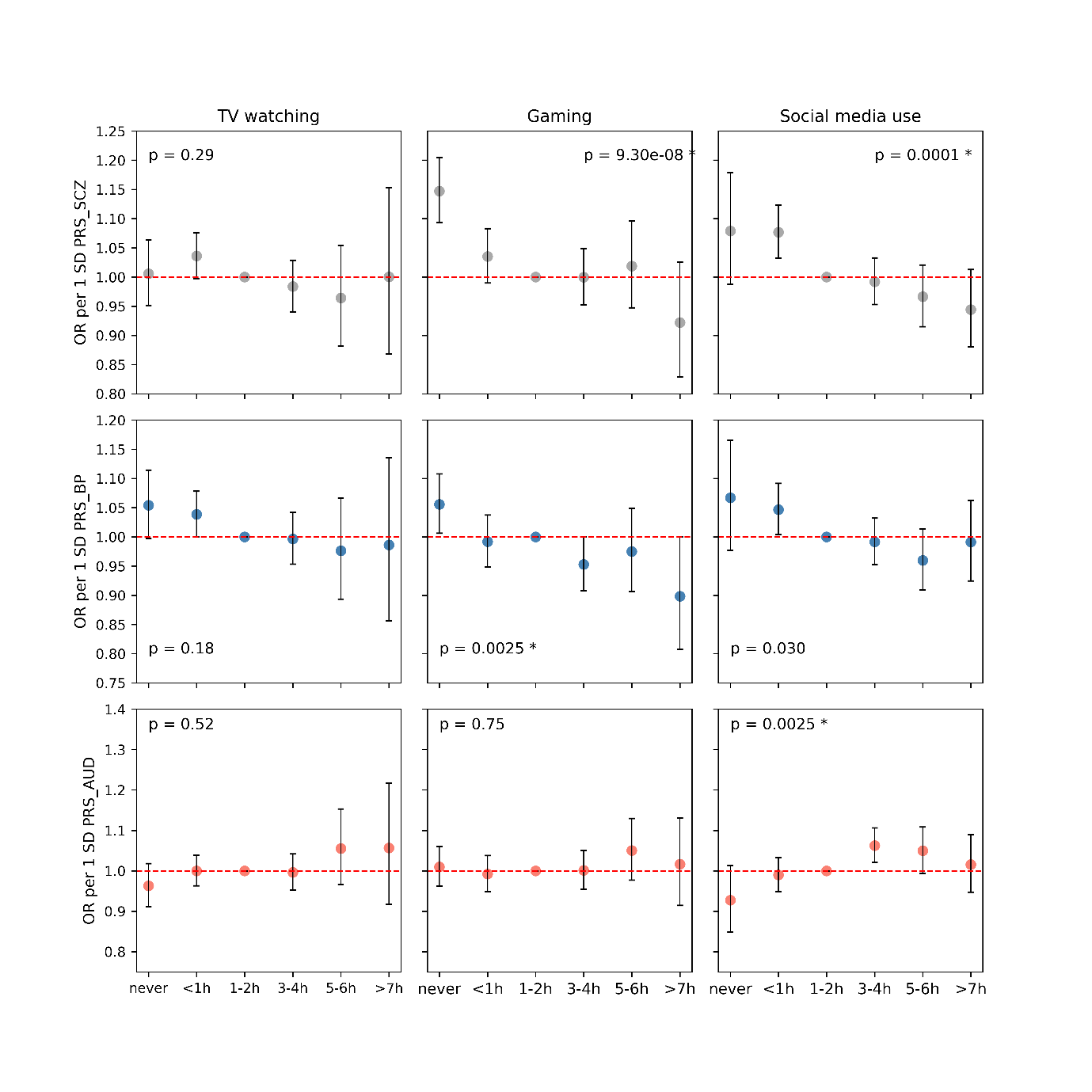

*Associations between levels of screen-based activities and polygenic risk scores were estimated using multinomial regression models, with screen use modeled as a categorical outcome variable with six levels (reference category set at “1–2 hours per day”). All models were adjusted for the first 10 genetic PCs and genotyping batch, and within each model age, genetic sex, and socioeconomic status were included as covariates. Analyses were performed only for diagnostic blocks/categories with a prevalence less than 1% in the study sample. P-values from the likelihood ratio tests comparing models with and without PRS are reported above each panel, with asterisks indicating statistically significant results after comparison to the family-wise error rate (α = 0.0056), adjusted using Bonferroni correction (n_tests_ = 9). The horizontal line indicates no association (OR = 1). Error bars represent 95% confidence intervals.*

*SCZ: schizophrenia; BP: bipolar disorder; AUD: alcohol use disorder; PRS: polygenic risk score*

### Figure S10. Associations between levels of screen-based activities and polygenic risk scores for major psychiatric disorders among participants without any psychiatric diagnoses registered (*n* = 14,625).

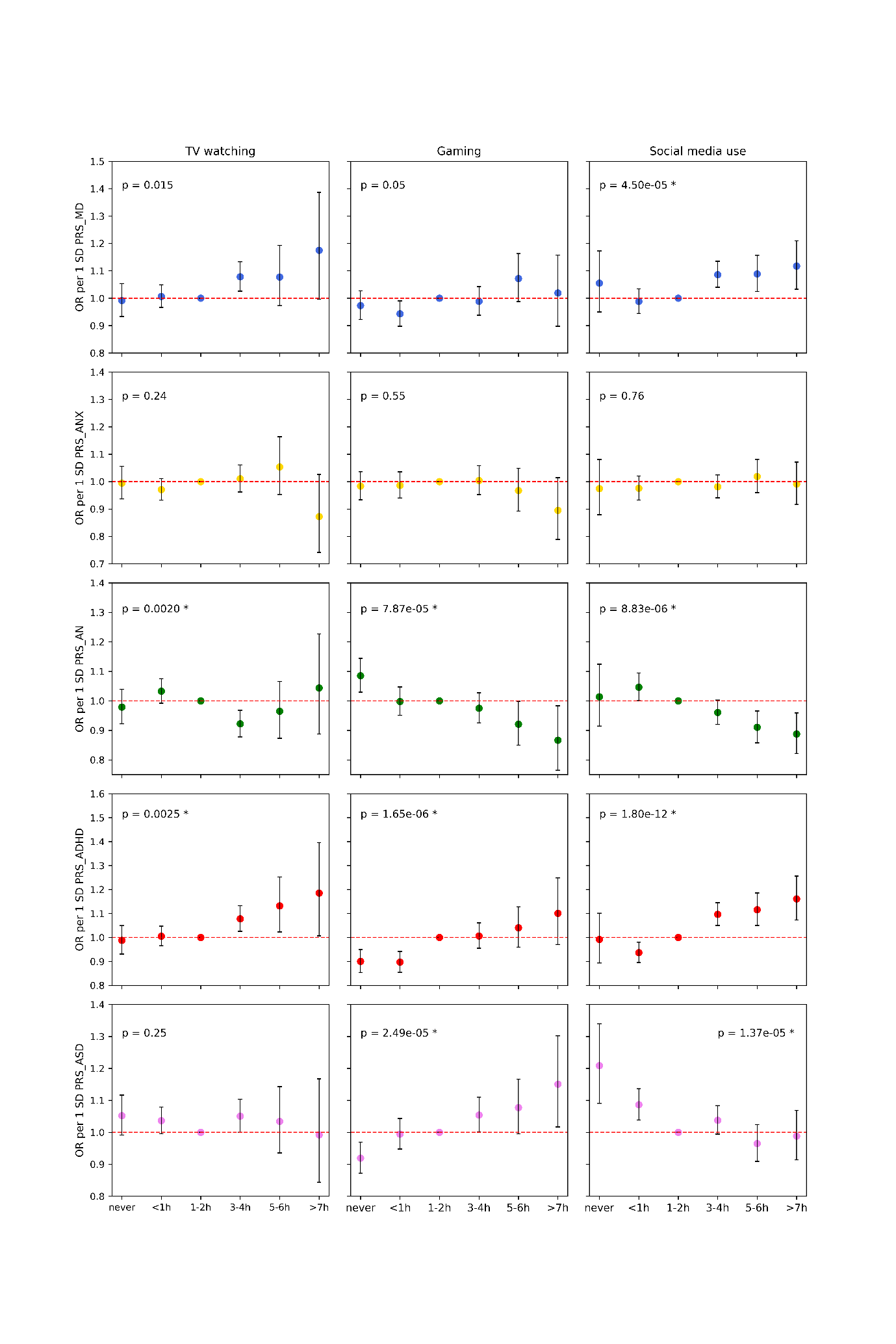

*Associations between levels of screen-based activities and polygenic risk scores for major psychiatric disorders were estimated using multinomial regression models, with screen use modeled as a categorical outcome variable with six levels (reference category set at “1–2 hours per day”). All models were adjusted for the first 10 genetic PCs and genotyping batch, and within each model age, genetic sex, and socioeconomic status were included as covariates. Analyses were performed only for diagnostic blocks/categories with a prevalence greater than 1% in the study sample. P-values from the likelihood ratio tests comparing models with and without PRS are reported above each panel, with asterisks indicating statistically significant results after comparison to the family-wise error rate (α = 0.0033), adjusted using Bonferroni correction (n_tests_ = 15). The horizontal line indicates no association (OR = 1). Error bars represent 95% confidence intervals.*

*MD: major depression; ANX: anxiety disorder; AN: anorexia nervosa; ADHD: attention-deficit hyperactivity disorder; ASD: autism spectrum disorder; PRS: polygenic risk score*

### Figure S11. Matrices of phenotypic correlations between polygenic risk scores, TV watching score, and diagnostic outcomes in the subsample with moderate-to-high TV watching level (*n* = 10,954), stratified by sex assigned at birth.

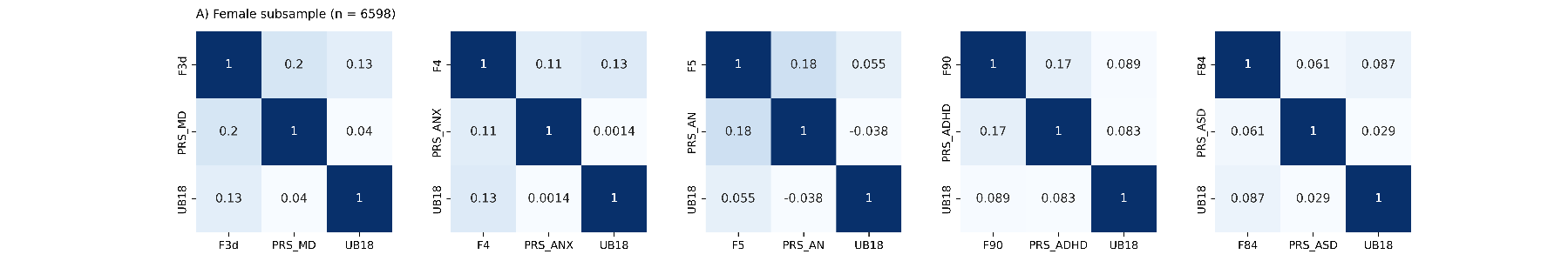

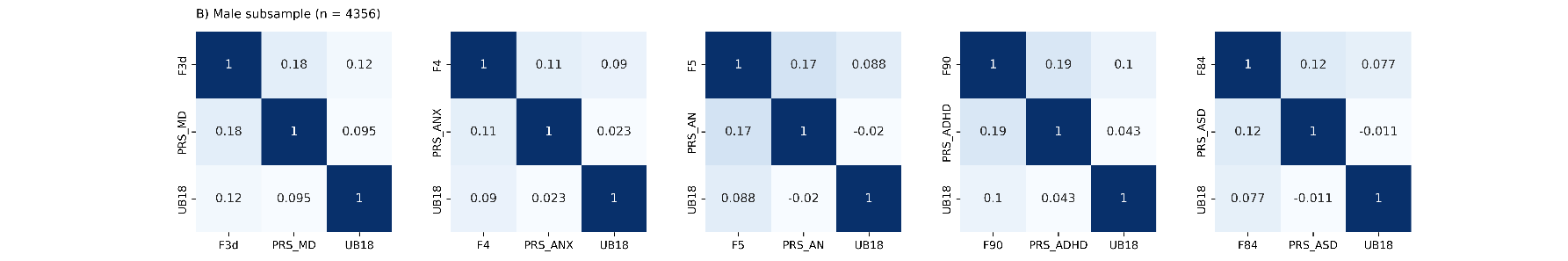

*Screen use level was classified as moderate-to-high if the reported score was 3 or above (i.e. 1–2 hours per day or more).*

*UB18: self-reported TV watching score; F3d: depressive disorder diagnosis; F4: anxiety/stress-related disorder diagnosis; F5: eating disorder diagnosis; F90: hyperkinetic disorder diagnosis; F84: pervasive developmental disorder diagnosis; PRS_MD: polygenic risk score for major depression; PRS_ANX: polygenic risk score for anxiety disorder; PRS_AN: polygenic risk score for anorexia nervosa; PRS_ADHD: polygenic risk score of attention-deficit hyperactivity disorder; PRS_ASD: polygenic score of autism spectrum disorder.*

### Figure S12. Matrices of phenotypic correlations between polygenic risk scores, TV watching score, and diagnostic outcomes in the subsample with moderate-to-high gaming level (*n* = 8541), stratified by sex assigned at birth.

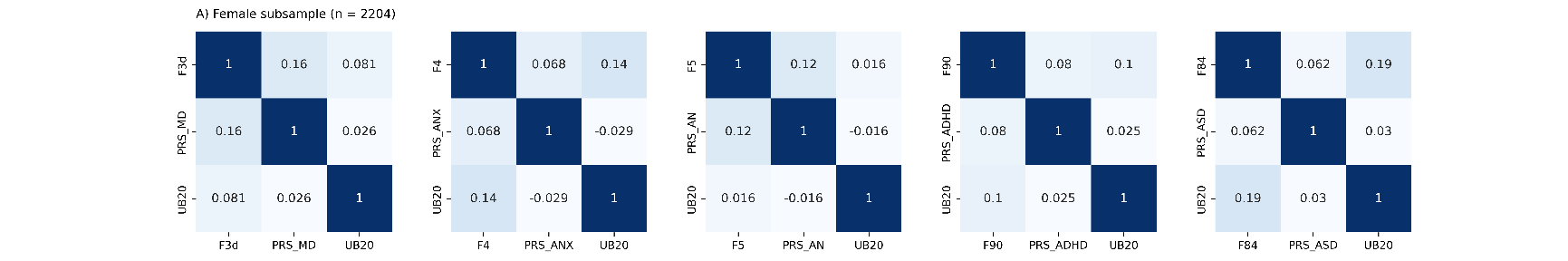

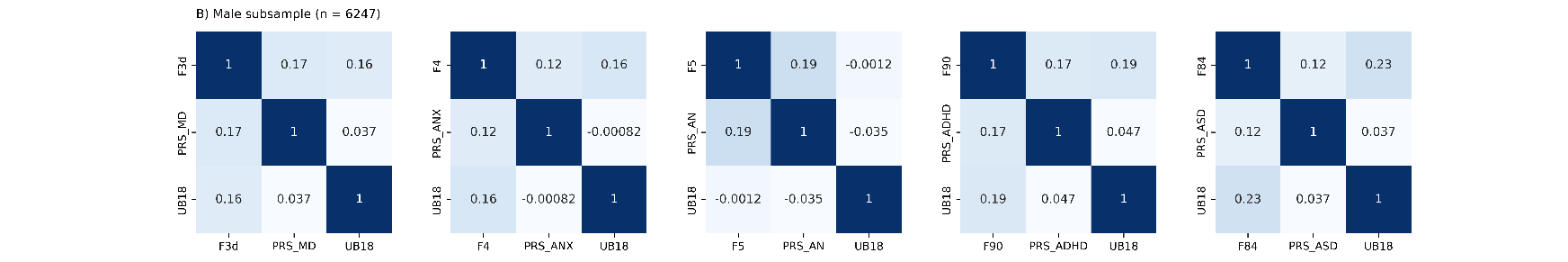

*Screen use level was classified as moderate-to-high if the reported score was 3 or above (i.e. 1–2 hours per day or more).*

*UB20: self-reported gaming score; F3d: depressive disorder diagnosis; F4: anxiety/stress-related disorder diagnosis; F5: eating disorder diagnosis; F90: hyperkinetic disorder diagnosis; F84: pervasive developmental disorder diagnosis; PRS_MD: polygenic risk score for major depression; PRS_ANX: polygenic risk score for anxiety disorder; PRS_AN: polygenic risk score for anorexia nervosa; PRS_ADHD: polygenic risk score of attention-deficit hyperactivity disorder; PRS_ASD: polygenic score of autism spectrum disorder.*

### Figure S13. Matrices of phenotypic correlations between polygenic risk scores, TV watching score, and diagnostic outcomes in the subsample with moderate-to-high social media use level (*n* = 12,821), stratified by sex assigned at birth.

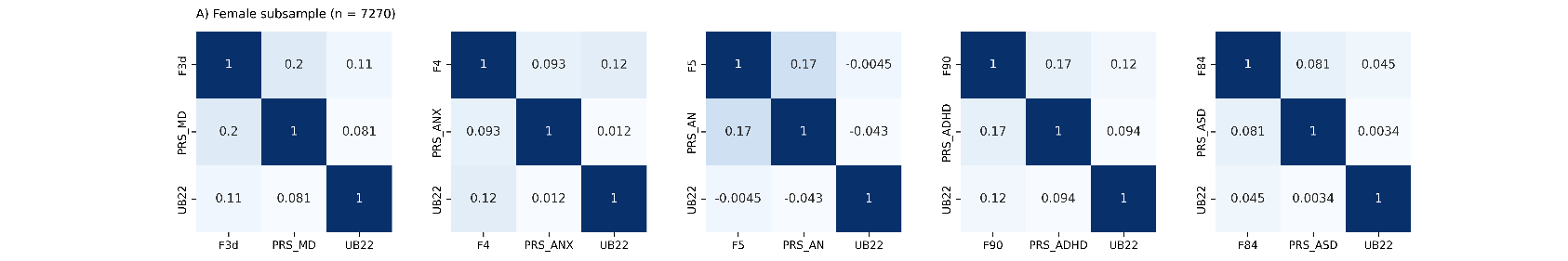

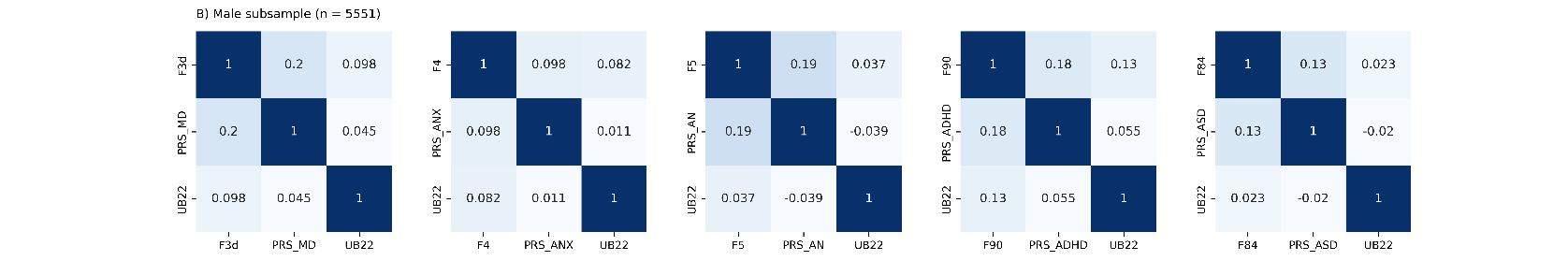

*Screen use level was classified as moderate-to-high if the reported score was 3 or above (i.e. 1–2 hours per day or more).*

*UB22: self-reported social media use score; F3d: depressive disorder diagnosis; F4: anxiety/stress-related disorder diagnosis; F5: eating disorder diagnosis; F90: hyperkinetic disorder diagnosis; F84: pervasive developmental disorder diagnosis; PRS_MD: polygenic risk score for major depression; PRS_ANX: polygenic risk score for anxiety disorder; PRS_AN: polygenic risk score for anorexia nervosa; PRS_ADHD: polygenic risk score of attention-deficit hyperactivity disorder; PRS_ASD: polygenic score of autism spectrum disorder.*

### Figure S14. Genetic confounding in the associations between moderate-to-high TV watching and diagnoses of depressive and hyperkinetic disorders, estimated separately for females and males.

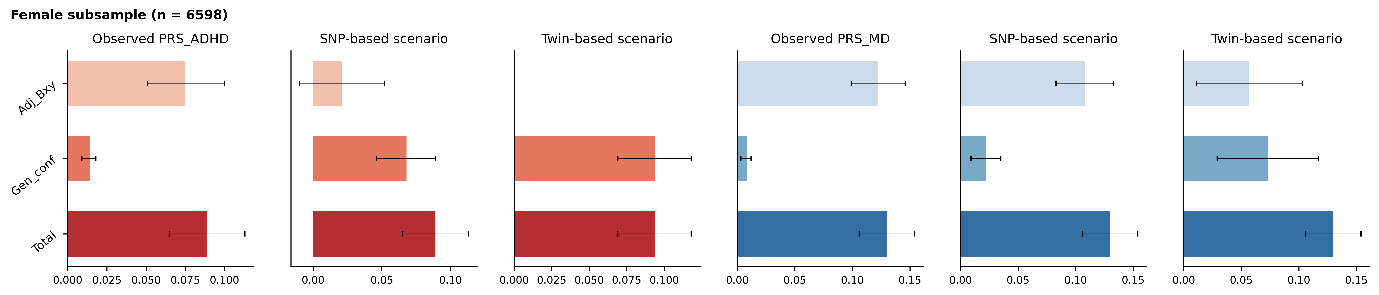

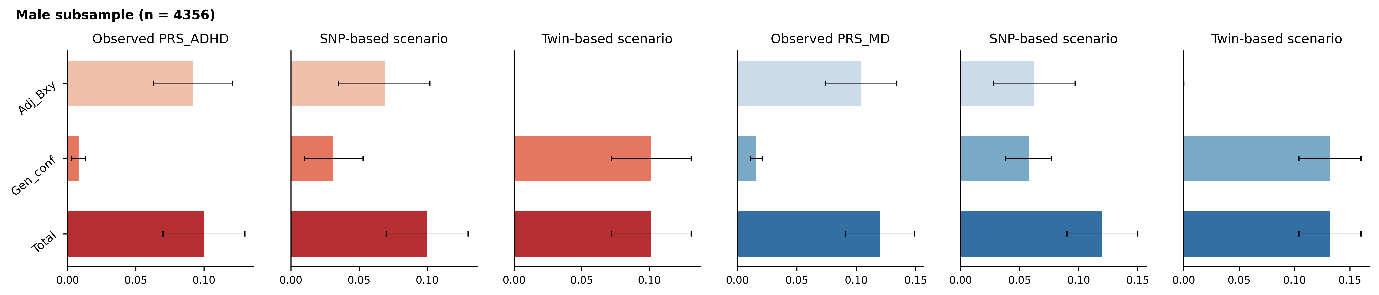

*Adj_Bxy: estimate of the relationship between moderate-to-high TV watching and diagnosis after accounting for the polygenic risk score (PRS) under three scenarios (observed PRS, PRS that explains SNP-heritability, and PRS that explains twin heritability); Gen_conf: estimate of genetic confounding; Total: total observed association.*

*PRS_MD: polygenic risk score for major depression; PRS_ADHD: polygenic risk score of attention-deficit hyperactivity disorder*

### Figure S15. Genetic confounding in the associations between moderate-to-high gaming and diagnoses of depressive, hyperkinetic, and pervasive developmental disorders, estimated separately for females and males.

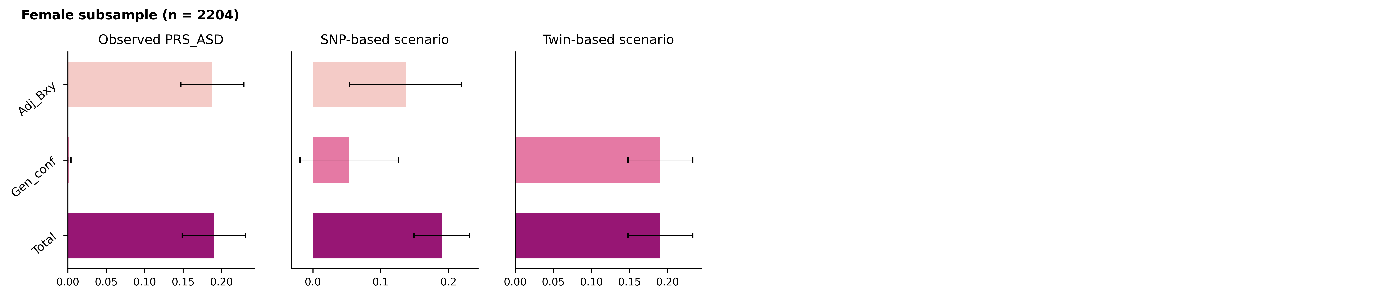

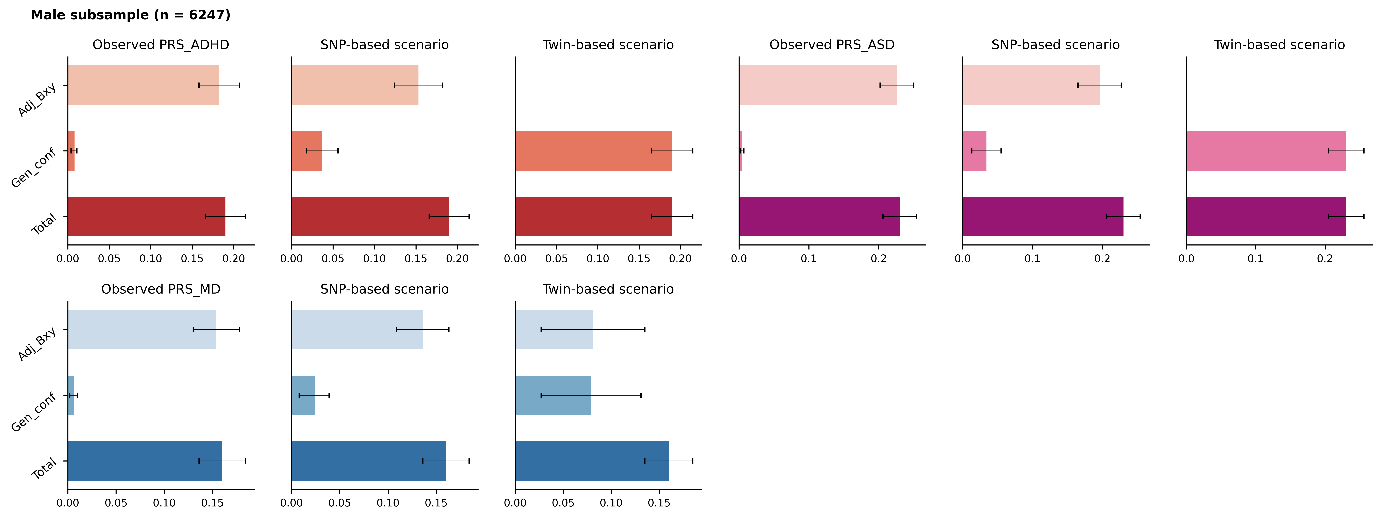

*Adj_Bxy: estimate of the relationship between moderate-to-high gaming and diagnosis after accounting for the polygenic risk score (PRS) under three scenarios (observed PRS, PRS that explains SNP-heritability, and PRS that explains twin heritability); Gen_conf: estimate of genetic confounding; Total: total observed association.*

*PRS_MD: polygenic risk score for major depression; PRS_ADHD: polygenic risk score of attention-deficit hyperactivity disorder; PRS_ASD: polygenic score of autism spectrum disorder.*

### Figure S16. Genetic confounding in the associations between moderate-to-high social media use and diagnoses of depressive and hyperkinetic disorders, estimated separately for females and males.

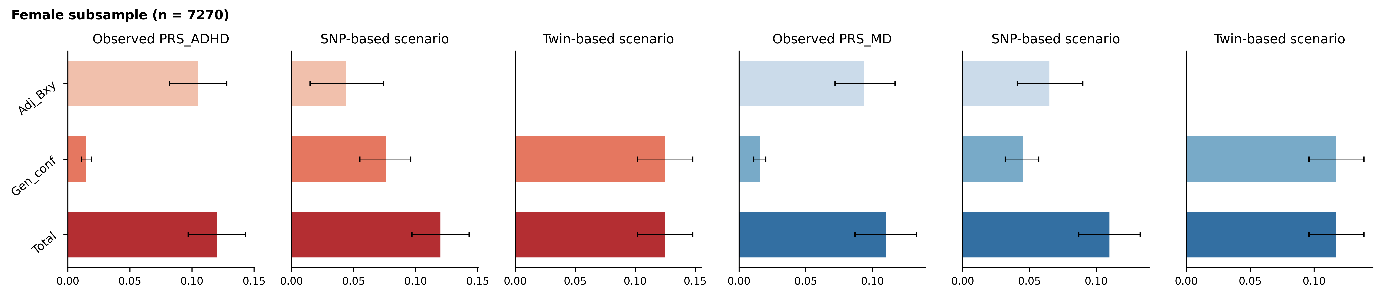

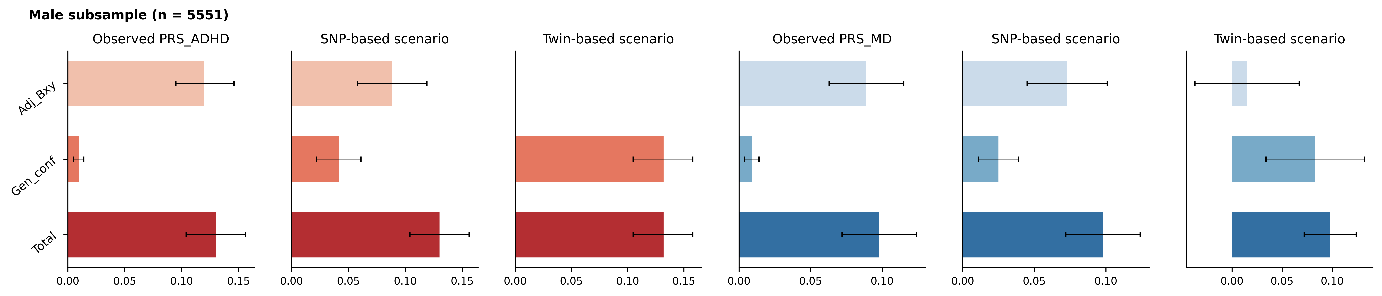

*Adj_Bxy: estimate of the relationship between moderate-to-high gaming and diagnosis after accounting for the polygenic risk score (PRS) under three scenarios (observed PRS, PRS that explains SNP-heritability, and PRS that explains twin heritability); Gen_conf: estimate of genetic confounding; Total: total observed association.*

*PRS_MD: polygenic risk score for major depression; PRS_ADHD: polygenic risk score of attention-deficit hyperactivity disorder*
